# A Smart Investment: The Health, Education, and Economic Returns of Malaria Chemoprevention in School-Aged Children Across Ten High-Burden Countries

**DOI:** 10.64898/2026.03.23.26349091

**Authors:** Katherine Snyman, Noam Angrist, Lauren M. Cohee, Eve Worall

## Abstract

Malaria imposes societal costs beyond health, including substantial effects on education, yet economic evaluations often overlook these broader impacts. We conducted a cross-sectoral benefit-cost analysis of malaria chemoprevention in school-aged children (SAC) across ten high-burden sub-Saharan African countries. Using recent trial data, we estimated impacts on malaria morbidity, mortality, school absenteeism, and literacy. The intervention was projected to cost $422 million and generate $5.7 billion in societal net benefits, yielding a benefit-cost ratio (BCR) of 14.3. Country-level BCRs ranged from 3.71 to 39.5, with the highest returns in Nigeria. Results were sensitive to drug choice, discount rate, and valuation of education benefits. When using school quality metrics (estimated via Learning-Adjusted Years of Schooling (LAYS)), BCRs increased up to 100-fold compared to estimates based on school quantity alone. Probabilistic sensitivity analysis yielded a mean simulated BCR of 11.00 (95% CI: 10.89–11.11), with a >95% probability of being cost-beneficial at a BCR threshold of 3. This study advances the evidence base for malaria chemoprevention in SAC, highlighting its dual health and educational benefits. These findings offer policymakers and funders strong evidence to prioritize malaria chemoprevention in SAC as a high-value investment in both health and human capital in malaria-endemic regions.

## 1 Introduction

Malaria remains a major public health challenge, with 263 million cases and 600,000 deaths annually (World Health Organization 2024). Two-thirds of the global malaria burden is concentrated in 11 African countries— Burkina Faso, Cameroon, the Democratic Republic of the Congo, Ghana, Mali, Mozambique, the Niger, Nigeria, the Sudan, Tanzania and Uganda—which are the focus of the WHO’s “High Burden to High Impact” (HBHI) initiative to accelerate malaria control (World Health Organization 2024). Substantial progress in malaria control was achieved between 2000 and 2015, particularly among children under five, through widespread distribution of insecticide-treated nets, deployment of seasonal malaria chemoprevention, and, more recently, the introduction of promising new vaccines (Bhatt et al. 2016; World Health Organization 2024). However, progress has stalled in recent years, with malaria cases and incidence per person at risk increasing between 2022 and 2023 (World Health Organization 2024).

School-aged children (SAC) (5–15 years old) represent a large, historically overlooked malaria risk group. An estimated 200 million SAC are at risk of malaria, with infection prevalence reaching as high as 50-80% in some settings (Nankabirwa et al. 2014; Brooker et al. 2017; Gething et al. 2011). In this group, malaria causes acute illness and chronic infection, anemia, and malaise, which impair cognitive function and physical well-being (Brooker et al. 2017). These effects can reduce educational achievement through increased absenteeism, impaired short-term cognition, and hindered long-term cognitive development, ultimately resulting in human capital losses (Fernando et al. 2006; Clarke et al. 2008; Angrist, Jukes, et al. 2023). Despite these negative impacts on both health and education, SAC remain largely overlooked in current malaria control strategies, exacerbating their relative disease burden. Although there are strong associations between malaria and reduced educational attainment (Barofsky, Anekwe, and Chase 2020; Lucas 2010; Kuecken, Thuilliez, and Valfort 2021), few randomized trials have quantified the human capital causal impacts of malaria on educational metrics, especially on education quality (in addition to education quantity).

Given stalled progress and budget constraints—where malaria funding is, at best, uncertain and, at worst, facing devastating cuts—there is an urgent need prioritize interventions that maximize both health and broader societal benefits (Burkybile 2025). Furthermore, school health has been identified as a key priority for achieving the Sustainable Development Goals (D. A. P. Bundy et al. 2017) and the WHO now officially recommends chemoprevention (the routine administration of full courses of anti-malarial drugs at regular intervals regardless of infection status to both clear existing infections and provide a period of prophylaxis against new infection) in SAC for malaria control in certain circumstances (World Health Organization 2023). Against this backdrop, we conducted the first benefit-cost analysis (BCA) of malaria chemoprevention in SAC, evaluating its potential to improve both health and education outcomes across ten HBHI countries.

This study makes two primary contributions. First, we estimate the economic impacts of malaria chemoprevention in SAC, using a conceptual framework (Figure 1) that incorporates health benefits (reduced malaria morbidity and mortality) and educational benefits (reduced absenteeism and improved literacy). Second, we advance the literature and methodology in malaria economics by jointly accounting for health gains and human capital accumulation, a dimension often missing from prior BCAs of malaria interventions (Shretta and Ngwafor Anye 2023; Korenromp et al. 2021; Lubell et al. 2008; Maitra, Hodge, and Jimenez Soto 2016).

**Figure 1.**
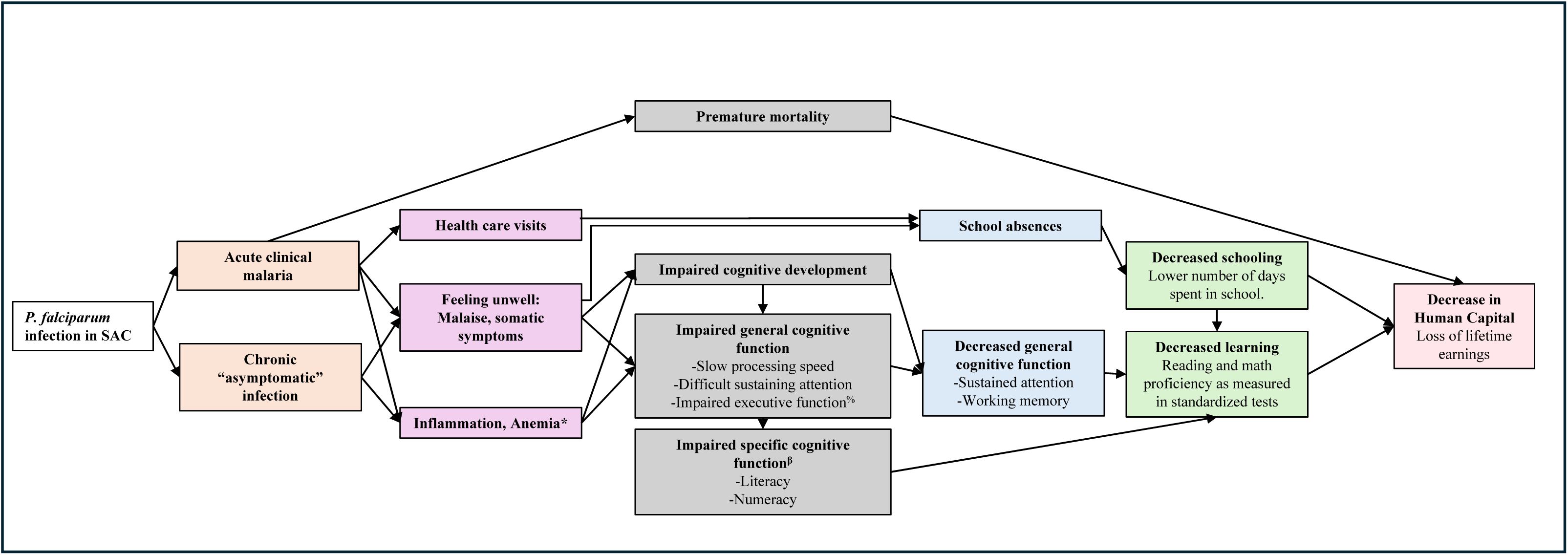
Conceptual framework the impact of malaria in school-age children. Caption: SAC: School-aged children; * P. falciparum is an independent cause of anemia, but also exacerbates other multifactorial causes of anemia, e.g. nutritional deficiencies and helminth infections. ^%^ Working memory, attention shifting/cognitive flexibility, inhibitory control, organization/planning ^β^ Reading: Phonological awareness, Decoding, Orthographic processing, Semantic processing; Math: Number sense, visual-spatial processing, pattern recognition, logical reasoning

**Figure 2.**
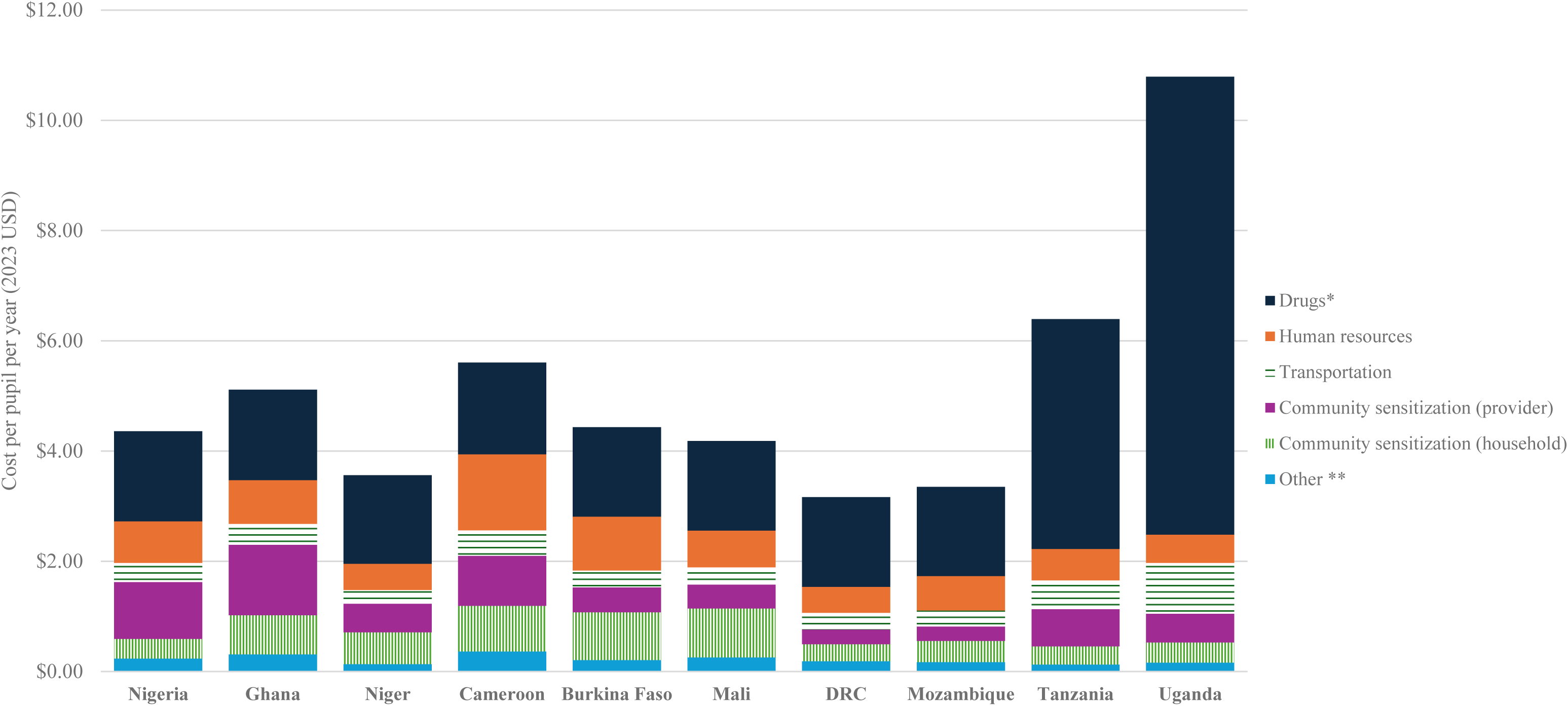
Annual intervention cost per pupil (societal perspective) Caption: All costs in 2023 USD. * Includes storage ** Includes supplies and materials and monitoring and evaluation

**Figure 3.**
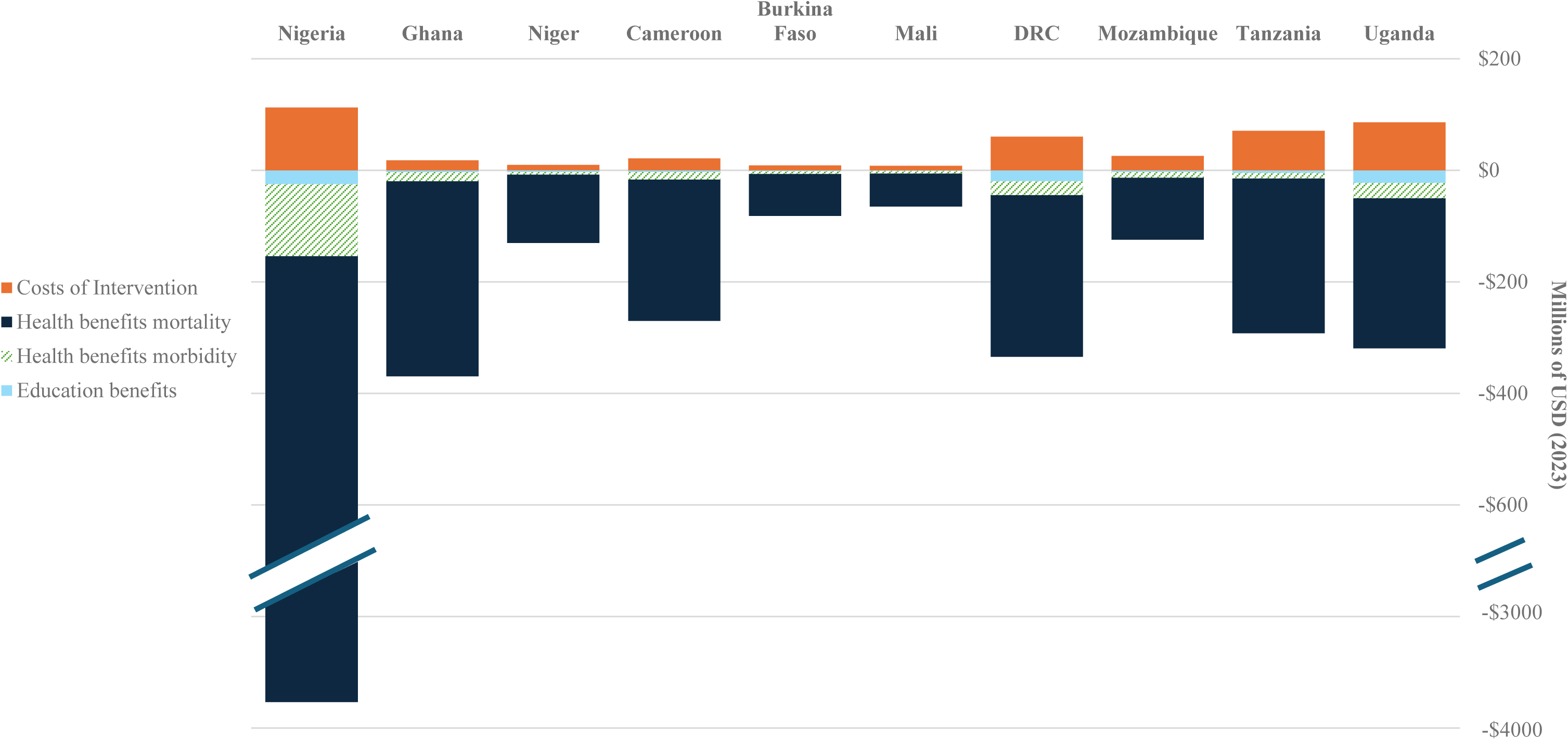
Incremental costs and benefits of malaria chemoprevention in SAC (societal perspective) All costs presented in 2023 USD. Educational gains estimated using the absenteeism-approach. A break in the y-axis is included to accommodate Nigeria’s substantially larger benefit values.

Although there is substantive literature demonstrating the cost-effectiveness of malaria interventions (White et al. 2011; Conteh et al. 2021), cost-effectiveness analysis misses certain potential benefits (and costs) and preclude comparison with other priority global investments, for example, in education, climate and economic development. Benefit-cost analyses are the appropriate economic evaluation method to compare across sectors, yet these remain relatively rare in malaria (n = 7 BCA studies in the past 15 years), with reported benefit-costs ratios (BCR) (range: <1 to 48) (Shretta and Ngwafor Anye 2023; Korenromp et al. 2021; Ezennia, Nduka, and Ekwunife 2017; Rajkumar, Guakler, and Tilahun 2012; Jamison et al. 2013; Mouzin et al. 2011; Purdy et al. 2013)(Supplementary Table S1). Human capital losses have not been incorporated in malaria-focused BCAs, except for Korenromp et al (2022), which quantified educational losses due to absenteeism. We advance this work by estimating education effects through two methods: first, education quantity alone and second, considering both education quantity and quality jointly, estimated via the learning-adjusted years of schooling (LAYS) framework aligned with the World Bank’s Human Capital Index (Angrist et al. 2020).

Our modeling results suggest that malaria chemoprevention in SAC in ten HBHI countries (excluding Sudan) could generate substantial net benefits, potentially averting an estimated 9.1 million malaria cases, 20,367 deaths, and 389,765 DALYs, with mortality risk reduction benefits accounting for most economic benefits. The aggregated societal BCR was 14.28, indicating that every $1 invested generated approximately $14 in benefits. Modelled benefits exceeded costs in all countries analyzed (BCR range: 3.71–39.5), particularly in countries where malaria burden, case fatality rates, and GNI per capita levels were higher. Probabilistic sensitivity analysis produced a mean simulated generalized societal BCR of 11.00 (95% CI: 10.89–11.11; range: 1.02-79.12), with a >95% probability of being cost-beneficial at a BCR threshold of 3.

Sensitivity analyses highlighted that assumptions about mortality valuation and education valuation significantly affect results; when considering school quality (e.g. using literacy measures) with the LAYS approach, this yields BCRs up to one-hundred times higher than school quantity (e.g. using absenteeism measures). Educational gains amplify health returns substantially: malaria chemoprevention in SAC led to 46.3 million additional school days attended and 18.5 million LAYS, placing it among the most cost-effective education interventions globally (World Bank 2020; Angrist et al. 2020). These results suggest that malaria chemoprevention in SAC is a highly attractive investment for malaria-endemic regions and underscore the value of cross-sectoral strategies that invest in human capital through integrated health and education interventions.

## 2 Methodology

### 2.1 Overview

Our methodology drew on benefit-cost analysis (Robinson et al. 2019) and cost-effectiveness analysis (Wilkinson et al. 2016) reference cases. We adopted a societal perspective, presenting combined costs to both the provider (government) and households, and used a microeconomic approach to estimate the intervention’s benefits and costs. First, we conducted a benefit-cost analysis for each country individually. Second, we combined the ten country-specific point estimates to generate an aggregate result across the ten HBHI countries. Third, we performed a simulated probabilistic generalized benefit-cost analysis and probabilistic sensitivity analysis, to model average conditions across the study settings and explore the impact of parameter variation and uncertainty on BCRs.

### 2.2 Conceptual framework

In addition to poor health outcomes, malaria is also linked to increased school absenteeism, impaired cognitive development and function, and, consequently, reduced learning outcomes. These factors limit the ability of SAC to reach their full potential, affecting both individual academic achievement and broader human capital development. The long-term economic consequences can be substantial. To illustrate these dynamics, we developed a conceptual framework (Figure 1), which highlights the interplay between malaria and education, at the individual pupil level. This framework was designed for settings with high malaria burden and is directly applicable to our study population.

### 2.3 Intervention

Malaria chemoprevention may be delivered to SAC, either as intermittent preventive treatment or extended-age seasonal malaria chemoprevention (Morlino et al. 2025). Both involve administering full courses of antimalarials at regular intervals, targeting age groups according to local malaria burden. Delivery can occur in communities or at fixed sites like schools. Our analysis modelled chemoprevention delivered to SAC enrolled in government-funded primary schools using a school-based delivery platform.

For the intervention drug, we assumed the use of sulfadoxine-pyrimethamine plus amodiaquine (SPAQ) in all countries, except Uganda and Tanzania. In these two countries, where either programmatic delivery is underway (Tanzania) or a clinical trial is planned (Uganda), we assumed the use of dihydroartemisinin-piperaquine (DP)(National Institute for Medical Research 2023). We assumed six rounds of treatment for all countries except Tanzania, where current practice is to administer malaria chemoprevention in SAC three times per year.

### 2.4 Decision tree model

We used a decision tree model to evaluate the health outcomes, educational outcomes, and associated costs of implementing malaria chemoprevention in SAC. The model estimated the costs of implementing malaria chemoprevention in SAC over one year, capturing associated morbidity-related cost savings within that timeframe, while mortality risk reduction and educational benefits were projected over the cohort’s lifespan. We applied a 5% discount rate consistently across both the benefit–cost and cost–effectiveness analyses, in line with the Copenhagen Consensus methodology and recent literature suggesting that 5% better reflects LMIC contexts than the 3% rate recommended in the iDSI reference case (Copenhagen Consensus Center, 2021; Haacker et al., 2019; Wilkinson et al., 2016).

Our analysis focused on ten African countries designated as HBHI priority countries^1^ by the World Health Organization (WHO), including Burkina Faso, Cameroon, the Democratic Republic of the Congo, Ghana, Mali, Mozambique, the Niger, Nigeria, Tanzania and Uganda (World Health Organization 2018).

### 2.5 Data

Population of children in government funded primary schools in 2024 came from the UNESCO indicator data of total primary school pupils and percentage enrolled at private schools and using sub-Saharan wide pupil growth rates (“UNESCO Institute for Statistics (UIS)” 2024). Data on primary school years and the number of primary schools per country were obtained from national sources, varying by country. Where available, country-specific inputs for costs, epidemiological data, and educational variables were used. Detailed data sources are provided below and in the parameters table (Supplementary Table S2).

## 3 Economic evaluation of health outcomes and avoided cases of death

All costs were inflated to 2023 country and year specific local currency units using gross domestic product deflators and then converted into 2023 USD using market exchange rates (World Bank 2025). All costs are presented in 2023 USD.

### 3.1 Intervention cost

We estimated the economic costs for both households and providers (government), categorizing costs into drugs, supplies and materials, human resources, community sensitization, storage, transportation, and monitoring and evaluation. Drug costs were sourced from Global Fund reference pricing, with an additional 5% for wastage and 10% for transportation, customs clearance, and insurance (Global Fund 2024b; 2024a; Montresor et al. 2010). We assumed 90% intervention coverage for children that are enrolled in government-funded primary schools (GiveWell 2021). For drug administration, we estimated that six teachers per primary school would each dedicate two hours per round (12 hours total) to administering the drugs to pupils. Additionally, we assumed that 50% of parents would attend a two-hour informational session, with time valued in relation to gross national income (GNI) per capita (World Bank 2025). Alternative approaches, including statutory minimum wages, informal wage rates, and household consumption data, were considered but not applied due to concerns regarding comparability between and data availability across the ten countries. Costs for training, community sensitization (including social behavior change campaigns), monitoring and evaluation, and administrative personnel were modelled drawing from costs analyses of interventions in Uganda and Kenya (Gonahasa et al. 2024; Zulaika et al. 2023). Across countries, costs varied by the number of pupils, schools and districts per country and by cost of drugs, teacher salaries and value per lost hour (GNI per capita). Consistent with our one-year analytic time frame, we excluded broader education system costs (e.g., teacher salaries or classroom expansion), as these are considered fixed in the short term and unlikely to be affected by the intervention within this period.

### 3.2 Estimating Health Outcomes

#### 3.2.1 Malaria cases and deaths

We obtained country-specific malaria case and death estimates from the *World Malaria Report 2024* and applied them to 2024 population estimates (World Health Organization 2024; United Nations 2024). To estimate malaria incidence among school-aged children, we assumed the proportion of malaria cases occurring in this age group based on Ugandan surveillance data (Namuganga et al. 2022). We then applied this proportion to each country’s total expected malaria cases and divided the result by the percentage of the population estimated to be in primary school (United Nations 2024).

The effect size of malaria chemoprevention in SAC was varied by drug choice and dosing frequency. For DP administered six times annually, we used a 50% reduction in clinical malaria (incidence rate ratio = 0.50; 95% CI: 0.39–0.60), based on a recent meta-analysis (Cohee et al. 2020). For DP administered three times per year, we assumed a 20% reduction (incidence rate ratio = 0.80; 95% CI: 0.71–0.91) in malaria incidence, using trial data from Makenga et al. (Makenga et al. 2023). In scenarios where SPAQ was administered six times per year, we assumed a proportionally lower protective effect compared to DP, reflecting its shorter duration of protection (4 weeks vs. 6 weeks)(Zongo et al. 2015). Accordingly, we applied two-thirds of the impact observed with DP, resulting in an estimated incidence rate ratio of 0.67. These effect sizes were used to estimate the total reduction in malaria cases associated with each chemoprevention regimen. The *World Malaria Report* estimates that 1–3% of malaria cases progress to severe disease; in our base case analysis, we applied a midpoint estimate of 2% to model this progression. Case fatality rates for all malaria cases were estimated using country-specific malaria deaths-to-cases ratios, which range from 0.13% to 0.44% (World Health Organization 2024).

#### 3.2.2 Disability-adjusted life years

To compare the cost-effectiveness of malaria chemoprevention in SAC with other malaria interventions, we estimated disability-adjusted life years (DALYs) using standard methods, no age weighting, and assuming deaths occur at age 9 for years of life lost calculations (World Health Organization 2020). Data inputs are detailed in Supplementary Materials (Table S2 & S4).

### 3.3 Valuing morbidity reductions

Using cost-of-illness methods, we modelled costs for three different categories of morbidity from symptomatic malaria cases/episodes: uncomplicated cases that did not seek care from a formal health provider; uncomplicated cases that sought care from a formal health provider; and severe malaria cases that required hospitalization (Jo 2014; Larg and Moss 2011).

#### 3.3.1 Direct savings to provider

For individuals that sought treatment at public health facilities, the provider incurred all costs related to drugs, diagnostics, and consultations. We assume all cases receive a malaria rapid diagnostic test and treatment with Artemether-Lumefantrine, costs from Global Fund reference pricing (Global Fund 2024b; The Global Fund 2023). The cost of outpatient consultations for uncomplicated malaria was estimated using WHO-CHOICE country-specific cost per outpatient visit (World Health Organization 2011). For inpatient cases, we estimated costs by multiplying the country-specific WHO-CHOICE cost per inpatient bed-day by the mean duration of hospitalization (3 days)(White et al. 2011; World Health Organization 2011).

#### 3.3.2 Direct savings to households

For those who sought treatment at private health facilities, we assumed that households incurred all cost related to drugs, test and consultation using the same methods described above. For all cases that sought care, we assumed that the household incurred additional out-of-pocket costs for transportation and food, based on estimates from Uganda (uncomplicated case: $0.50; severe case: $1.00)(Snyman et al. 2025).

#### 3.3.3 Indirect benefits to society

We assumed that untreated and treated malaria cases incurred productivity losses associated with time spent away from work due to caregiver time. Productivity losses were valued using the human capital approach (Hodgson and Meiners 1982; Hansen and Yeung 2009), where annual GNI per capita was divided by 365 and then multiplied by an assumed number of caregiver days lost for untreated and uncomplicated malaria (2 days)(Sicuri et al. 2011; Ayieko et al. 2009; Chima, Goodman, and Mills 2003; Snyman et al. 2025), and severe malaria (5 days)(White et al. 2011). We selected GNI per capita to ensure a standardized approach across countries. We did not quantify productivity losses for school-age children.

### 3.4 Valuing mortality risk reduction

We translated the estimated reduction in deaths into monetary benefits by multiplying deaths averted by the value of a statistical life (VSL). VSL represents a population’s aggregate willingness to pay for small reductions in mortality risk and is empirically derived from individuals’ willingness to trade income for marginal changes in their own risk of death. Because most primary VSL studies are conducted in high income settings, we extrapolated VSL estimates to each study country following the Reference Case (Robinson et al. 2019; Pradhan and Jamison 2019). Study country specific VSL were calculated as:

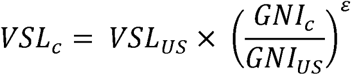

where VSL_US_ was the U.S. reference value, GNI_c_ was a study country-specific GNI per capita, GNI_US_ was the U.S. GNI per capita, and ε was the income elasticity of VSL. Income elasticity measures the responsiveness of a change in demand (in this case willingness to pay for reductions in mortality risk) to a change income, when elasticity is 1, the change is proportionate, and where its greater than 1, demand is highly responsive to income changes. Thus when ε ≠ 1, the VSL to income ratio differed across countries.

The base case used the Reference Case featured specification, applying an income elasticity of 1.5 to extrapolate the U.S. VSL to lower income settings. This higher elasticity reflects evidence that willingness to pay for mortality risk reductions increases more than proportionally with income, implying lower VSL-to-income ratios in lower-income settings. (Robinson et al. 2019). Sensitivity analyses followed the standardized Reference Case approach, assuming an elasticity of 1 and applying constant VSL-to-income ratios of 100 and 160 times GNI per capita. These ratios reflect values commonly observed in high-income settings and are recommended to facilitate comparison across studies and to explore uncertainty in extrapolated VSL estimates.

Because malaria deaths in our model occurred at approximately age 9, we derived an age specific VSL using a value per statistical life year (VSLY) approach. VSLY was calculated as:

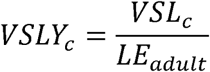

where LE_adult_ represented undiscounted remaining life expectancy at the average adult age. Age specific VSL was then calculated as:

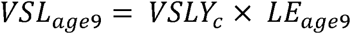

where LE_age9_ represented remaining life expectancy at age 9 based on country life tables (World Health Organization 2025). All costs and deaths averted occurred within a single program year. Because VSL represented a present-value measure of mortality risk reduction, no additional intertemporal discounting was applied. Detailed country-specific calculations are provided in the Supplementary Materials (Table S3).

## 4 Learning outcomes and economic impact

We used two approaches to model the educational gains from malaria chemoprevention in SAC and their economic impact.

### 4.1 Education Quantity as measured by Absenteeism

The base case approach used absenteeism reductions associated with malaria chemoprevention in SAC to estimate gains in earnings. We assumed a linear relationship between attendance and schooling progression, whereby each additional day of school contributes proportionally to overall grade progression. Using estimates of the average number of school days missed due to malaria (uncomplicated case: 5 days; severe case: 10 days) (Korenromp et al. 2021) and modelled estimates of cases averted through malaria chemoprevention in SAC, we calculated the total additional days of schooling gained per child annually. These additional days were then converted into fractions of a school year to estimate total years of schooling gained. To estimate the economic impact, we applied country-specific returns to schooling from the Mincer earnings function – an econometric model that predicts earnings based on education – to to translate these gains into increased expected lifetime earnings (Fink et al. 2016). Earnings were projected across the working lifespan (ages 15–60), discounted to present value, and adjusted for expected wage growth.

### 4.2 Combining Education Quantity and Quality measured by Foundational Skills in Literacy

The second approach translated foundational skills in literacy gains from an intermittent preventive treatment in school children trial in Malawi (Sixpence et al. 2024) into gains in LAYS and then into lifetime earnings. We calculated a Cohen’s *d* effect size (Baird and Pane 2019) from observed test score changes and sample size, then converted this into learning-adjusted years of schooling (LAYS) consistent with the micro-LAYS approach and Human Capital Index standards (Evans and Yuan 2019; Angrist et al. 2020). LAYS were then mapped to lifetime earnings gains using country-specific Mincerian returns (Fink et al. 2016) applied to discounted expected earnings from age 15–65, adjusting for wage growth.

## 5 Benefit-Cost and Cost-effectiveness Analyses

### 5.1 Benefit-Cost analysis

The modeled benefits of the malaria chemoprevention in SAC intervention included direct cost savings to the health system and households from reduced use of outpatient and inpatient services, indirect savings from reduced morbidity and mortality, and human capital gains from improved educational outcomes. These benefits were estimated using the outputs from the decision tree model, which provided estimates of malaria cases and deaths averted and educational gains in the malaria chemoprevention in SAC scenario relative to baseline.

We calculated the BCR, which quantifies the economic return per dollar spent, by dividing the net present value of the incremental health and educational benefits by the net present value of the incremental intervention costs. The Copenhagen Consensus “Traffic Light” rating system, which categorizes the BCR into four categories: Excellent (BCR ≥ 20), Good (5 ≤ BCR < 20), Fair (1 ≤ BCR < 5), and Poor (BCR < 1)(Angrist, Aurino, et al. 2023), was applied to aid interpretation of results. Additionally, the net benefits (total benefits minus total costs) are reported, as relying solely on the BCR can obscure the relative magnitude of the effects.

### 5.2 Sensitivity analysis

To address uncertainty in both model parameters and model structure, we performed several sensitivity analyses. First, in line with suggestions for Standardized Sensitivity Analysis in BCA (Robinson et al. 2019; Pradhan and Jamison 2019), we performed structural sensitivity analyses, testing different model assumptions for alternative discount rates and varying how educational benefits (reductions in absenteeism vs gains in LAYS due to foundational skills in literacy gains) and health benefits (VSL estimation methods) are monetized. Structural sensitivity analyses were performed for the individual country BCRs.

Second, we conducted a probabilistic sensitivity analysis using XLRisk (Vose Software 2021) to model average conditions across the ten HBHI countries and to assess the impact of parameter variation and uncertainty in the results. Input parameters were derived by taking the mean of fixed values across the 10 HBHI countries or fitting probability distributions to pooled data where appropriate (Supplementary Table S4). This approach produced a simulated country representing average conditions across the study settings. Monte Carlo simulations were used to vary all uncertain parameters across plausible distributions, generating a distribution of benefit-cost outcomes that reflected uncertainty and heterogeneity across the ten countries. The reported confidence interval reflects the precision of the simulated mean estimate given the number of iterations, whereas the full range of possible outcomes is represented by the percentile distribution. From this analysis, we derived a benefit-cost acceptability curve and produced a tornado diagram to illustrate the correlation between the BCR and the sampled input values, using the absolute values of these correlations to highlight influential parameters.

### 5.3 Cost-effectiveness analysis

We modelled cost-effectiveness over a 1-year intervention period, adopting a provider perspective for both intervention costs and cost-savings. Mortality risk reduction benefits and educational gains were not included. Results are presented as incremental cost-effectiveness ratios and compared against cost-effectiveness thresholds based on opportunity cost approaches, which better reflect the health gains that could be achieved if resources were allocated to other competing interventions in LMICs (Pichon-Riviere et al. 2023).

### 5.4 Limitations

#### 5.4.1 Malaria transmission and modelling assumptions

Firstly, we did not account for seasonality in malaria transmission in relation to the timing of school attendance, which may vary across countries and affect the impact of the intervention. In highly seasonal, high-burden areas, existing seasonal malaria chemoprevention programs targeting younger children may already be in place, and delivery to older children could be integrated through community-based channels even when schools are closed. Secondly, our analysis was limited to the direct benefits experienced by school-aged children receiving chemoprevention—namely, reductions in malaria cases and deaths and improved educational outcomes. However, school-aged children are known reservoirs of malaria transmission, and reducing infection in this group may also lower community-wide transmission. As the magnitude of this indirect effect is not yet well quantified in community-based studies, we excluded it from our analysis, likely underestimating total benefits and producing conservative BCRs. We valued time losses using GNI per capita to ensure comparability across countries,This is an imperfect proxy for opportunity cost, particularly in predominantly rural and informal labor markets where typical earnings may be lower than national averages. Alternative approaches, including statutory minimum wages, informal daily wage rates, or household consumption data, may better reflect local opportunity costs but were not consistently available across all ten countries (Setiawan et al., 2024).. To the extent that GNI per capita exceeds prevailing earnings, productivity losses and therefore net benefits and benefit cost ratios may be modestly overstated. However, productivity effects represent a minority share of total monetized benefits, and sensitivity analyses using lower time values did not materially alter our benefit-cost ratios.

#### 5.4.2 Educational impact estimation

Educational benefits were restricted to individual-level outcomes and did not account for classroom-level spillovers or broader societal and macroeconomic gains. In addition, we did not capture potential network or general equilibrium effects of long-term improvements in schooling and human capital formation, which likely means our BCR and ICER estimates are conservative. We used two approaches to estimate these benefits, each with strengths and limitations.

Calculating LAYS from standardized assessments of foundational skills in literacy/numeracy is a relatively new approach, but we believe it has promise, as it includes a learning adjustment that enables cross-country comparisons (Angrist et al. 2020). However, due to lack of evidence, our effect size was drawn from a single trial in Malawi (Sixpence et al. 2024). Recent meta-analyses concluded that while there are gains to some cognitive skills such as attention, there was no statistically significant observed increase in literacy or numeracy test scores in the broader literature (Angrist, Jukes, et al. 2023). However, there are few studies rigorously measuring learning. Further studies using standardized metrics for measuring foundational skills in literacy and math are needed. The absenteeism-based approach is supported by more consistent evidence, as the number of school days lost due to malaria is well documented (Halliday et al. 2020; Korenromp et al. 2021). However, this method assumes that rates of return to education, which are typically calculated based on educational attainment (i.e., additional years of schooling), can be applied linearly to smaller units such as months or, in our case, days—a relationship that has not been validated (Mincer 1974; Patrinos and Psacharopoulos 2020; Fink et al. 2016). Therefore, if more data were available, we would consider the foundational skills in literacy LAYS approach preferable; however, there is currently a weaker evidence base.

#### 5.4.3 Equity and social protection considerations

We do not disaggregate our results by equity-relevant variables such as gender or household wealth, despite known variation in key inputs like school enrollment (UNESCO Institute for Statistics 2020), health-seeking behavior (Galactionova et al. 2017), and malaria transmission intensity (Tusting et al. 2013). These differences may influence both the effectiveness and cost-effectiveness of the intervention. Moreover, while school-based delivery can be efficient, it may overlook out-of-school children who are often more disadvantaged. Gender-specific benefit-cost analyses may also yield different results; for example, female education is linked to broader health and economic gains, including reductions in child and adult mortality (Pradhan et al. 2017). Finally, we did not evaluate the role of malaria chemoprevention in promoting social protection or mitigating household economic shocks. Future research should aim to capture these broader impacts and assess how malaria chemoprevention can support more equitable outcomes.

#### 5.4.4 Economies of scope

Our analysis did not account for potential economies of scope that could be achieved by integrating chemoprevention for SAC with existing school-based health interventions, such as deworming or nutrition programs, and with malaria control measures such as ITN distribution. Leveraging existing delivery platforms for SAC could reduce delivery costs and create synergies that further strengthen the value for money of this intervention.

## 6 Findings

### 6.1 Cost projections

Our model found that the aggregate cost of implementing malaria chemoprevention in SAC for one year in the ten HBHI countries was $422 million, with 13% of total costs incurred by households. The modeled societal annual cost of malaria chemoprevention in SAC delivery ranged from $9 million to $113 million per country (Figure 1, Table 1, Supplementary Table S5). Drug costs were the primary cost driver, accounting for 68–81% of total societal costs in countries using DP, compared to 30–53% in SPAQ-using countries. The cost per dose delivered (excluding drug costs) was similar across countries, ranging from $0.28 in Niger to $0.67 in Tanzania, with the main cost drivers being mean number of pupils per primary school and number of doses administered (3 vs. 6). These values align with published estimates of the cost per dose delivered (excluding drugs) for school-based deworming programs in Kenya (GiveWell 2021) and Vietnam (Caesar delos Trinos et al. 2023).

**Table 1.** Incremental costs, health and educational benefits and benefit-cost ratios. All costs presented in millions 2023 USD. Educational gains estimated using the absenteeism-approach. Green color: “excellent” rating; yellow color: “good” rating; orange: “fair rating” according to the Copenhagen Consensus ‘traffic light’ rating system. DRC: Democratic Republic of the Congo; HBHI: High Burden High Impact; DP: Dihydroartemisinin-piperaquine; SPAQ: Sulfadoxine-pyrimethamine + Amodiaquine.

|  | Nigeria | Ghana | Niger | Cameroon | Burkina Faso | Mali | DRC | Mozambique | Tanzania | Uganda | Aggregate<br>HBHI<br>Countries |
| --- | --- | --- | --- | --- | --- | --- | --- | --- | --- | --- | --- |
| **Intervention Drug & # of doses** | SPAQ X6 | SPAQ X6 | SPAQ X6 | SPAQ X6 | SPAQ X6 | SPAQ X6 | SPAQ X6 | SPAQ X6 | DP X3 | DP X6 |  |
| **Intervention Cost (millions)** | \$112.87 | \$18.02 | \$9.87 | \$21.49 | \$8.99 | \$8.20 | \$60.57 | \$25.88 | \$70.94 | \$86.12 | \$422.97 |
| **Incremental Malaria Cases averted** | 2,995,407 | 297,961 | 337,742 | 386,411 | 265,002 | 256,515 | 2,169,909 | 825,922 | 36,6632 | 1,217,059 | 9,118,561 |
| **Incremental Deaths averted** | 8,119 | 521 | 1,497 | 611 | 526 | 443 | 4,417 | 1,595 | 1,095 | 1,544 | 20,367 |
| **Incremental Morbidity Cost Savings (millions)** | \$128.99 | \$16.21 | \$4.30 | \$13.23 | \$4.81 | \$4.59 | \$25.07 | \$10.07 | \$10.04 | \$27.81 | \$245.11 |
| **Incremental Mortality Cost Savings (millions)** | \$3,900.58 | \$349.94 | \$122.52 | \$253.56 | \$75.37 | \$59.06 | \$289.99 | \$111.08 | \$277.65 | \$269.08 | \$5,708.83 |
| **Net Benefits (health only) (millions)** | \$3,916.70 | \$348.13 | \$116.95 | \$245.30 | \$71.19 | \$55.44 | \$254.49 | \$95.27 | \$216.75 | \$210.77 | \$5,530.97 |
| **BCR (health only)** | 35.70 | 20.32 | 12.84 | 12.41 | 8.92 | 7.76 | 5.20 | 4.68 | 4.06 | 3.45 | 13.08 |
| **Incremental school years gained** | 41631 | 4149 | 4678 | 5382 | 3688 | 3572 | 30198 | 11497 | 5093 | 16963 | 126,852 |
| **Incremental Human Capital Gains due to Education (millions)** | \$24.69 | \$3.39 | \$3.24 | \$3.17 | \$1.54 | \$1.16 | \$19.41 | \$3.16 | \$4.54 | \$22.41 | \$86.72 |
| **Net Benefits (health and education) (millions)** | \$3,941.39 | \$351.51 | \$120.19 | \$248.48 | \$72.73 | \$56.61 | \$273.90 | \$98.43 | \$221.29 | \$233.18 | \$5,617.70 |
| **BCR (health and education)** | 35.92 | 20.50 | 13.17 | 12.56 | 9.09 | 7.90 | 5.52 | 4.80 | 4.12 | 3.71 | 14.28 |

**Table 2.** Structural Sensitivity Analysis for discount rate, method of valuing mortality-risk reduction benefits and education benefits. Benefit-cost ratios (BCRs) estimated using the societal perspective. Countries ordered from most cost-beneficial to least. *Indicates base case. Green color: “excellent” rating; yellow color: “good” rating; orange: “fair” rating according to the Copenhagen Consensus ‘traffic light’ rating system. ; DP: Dihydroartemisinin-piperaquine; LAYS: Learning-adjusted school years; SPAQ: Sulfadoxine-pyrimethamine + Amodiaquine. VSL: Value per statistical life; VSLY: Value per statistical life year

| Country<br>*Drug x doses per year* | Mortality<br>valuation<br>method | Education Outcome Method |  |  |  |  |  |
| --- | --- | --- | --- | --- | --- | --- | --- |
|  |  | Absenteeism method |  |  | Foundational skill in literacy LAYS method |  |  |
|  |  | BCR (8% discount rate) | BCR (5% discount rate*) | BCR (1% discount rate) | BCR (8% discount rate) | BCR (5% discount rate*) | BCR (1% discount rate) |
| **Nigeria**<br>*SPAQ x 6* | VSL Y* | 35.8 | 35.9 | 36.3 | 188.1 | 311.8 | 776.1 |
|  | VSL SA1 | 20.0 | 20.1 | 20.6 | 172.3 | 296.0 | 760.4 |
|  | VSL SA2 | 44.6 | 44.7 | 45.1 | 196.9 | 320.6 | 784.9 |
|  | VSL SA3 | 70.5 | 70.6 | 71.1 | 222.9 | 346.6 | 810.9 |
| **Ghana**<br>*SPAQ x 6* | VSL Y* | 20.4 | 20.5 | 20.9 | 230.8 | 405.5 | 1,068.7 |
|  | VSL SA1 | 11.4 | 11.5 | 11.9 | 221.8 | 396.5 | 1,059.7 |
|  | VSL SA2 | 22.6 | 22.6 | 23.0 | 233.0 | 407.6 | 1,070.9 |
|  | VSL SA3 | 35.5 | 35.6 | 35.9 | 245.9 | 420.6 | 1,083.8 |
| **Niger**<br>*SPAQ x 6* | VSL Y* | 13.0 | 13.2 | 13.8 | 69.5 | 114.4 | 281.1 |
|  | VSL SA1 | 7.2 | 7.3 | 8.0 | 63.6 | 108.6 | 275.3 |
|  | VSL SA2 | 28.2 | 28.3 | 29.0 | 84.6 | 129.6 | 296.3 |
|  | VSL SA3 | 44.7 | 44.9 | 45.5 | 101.2 | 146.1 | 312.8 |
| **Cameroon**<br>*SPAQ x 6* | VSL Y* | 12.5 | 12.6 | 12.8 | 121.3 | 199.3 | 472.9 |
|  | VSL SA1 | 6.8 | 6.9 | 7.1 | 115.7 | 193.6 | 467.2 |
|  | VSL SA2 | 15.8 | 15.8 | 16.1 | 124.6 | 202.6 | 476.1 |
|  | VSL SA3 | 24.8 | 24.9 | 25.1 | 133.6 | 211.6 | 485.2 |
| **Burkina Faso**<br>*SPAQ x 6* | VSL Y* | 9.0 | 9.1 | 9.4 | 80.3 | 139.6 | 364.5 |
|  | VSL SA1 | 5.1 | 5.1 | 5.5 | 76.4 | 135.6 | 360.6 |
|  | VSL SA2 | 16.1 | 16.1 | 16.5 | 87.4 | 146.6 | 371.6 |
|  | VSL SA3 | 25.3 | 25.4 | 25.7 | 96.6 | 155.9 | 380.8 |
| **Mali**<br>*SPAQ x 6* | VSL Y* | 7.8 | 7.9 | 8.1 | 56.4 | 90.6 | 210.0 |
|  | VSL SA1 | 4.5 | 4.5 | 4.8 | 53.0 | 87.3 | 206.6 |
|  | VSL SA2 | 14.3 | 14.4 | 14.6 | 62.8 | 97.1 | 216.4 |
|  | VSL SA3 | 22.5 | 22.5 | 22.8 | 71.0 | 105.3 | 224.6 |
| **Democratic Republic of the Congo**<br>*SPAQ x 6* | VSL Y* | 5.3 | 5.5 | 6.4 | 168.3 | 342.2 | 1,087.4 |
|  | VSL SA1 | 3.1 | 3.2 | 4.1 | 166.0 | 339.9 | 1,085.1 |
|  | VSL SA2 | 11.9 | 12.1 | 13.0 | 174.9 | 348.8 | 1,094.0 |
|  | VSL SA3 | 18.8 | 18.9 | 19.8 | 181.7 | 355.6 | 1,100.8 |
| **Mozambique**<br>*SPAQ x 6* | VSL Y* | 4.8 | 4.8 | 5.0 | 80.1 | 132.6 | 313.6 |
|  | VSL SA1 | 2.5 | 2.6 | 2.8 | 77.9 | 130.3 | 311.4 |
|  | VSL SA2 | 9.9 | 9.9 | 10.1 | 85.2 | 137.7 | 318.7 |
|  | VSL SA3 | 15.5 | 15.6 | 15.8 | 90.9 | 143.3 | 324.4 |
| **Tanzania**<br>*DP x 3* | VSL Y* | 4.1 | 4.1 | 4.2 | 119.2 | 206.6 | 523.4 |
|  | VSL SA1 | 2.3 | 2.3 | 2.5 | 117.4 | 204.8 | 521.6 |
|  | VSL SA2 | 6.3 | 6.3 | 6.4 | 121.4 | 208.8 | 525.6 |
|  | VSL SA3 | 9.9 | 9.9 | 10.0 | 125.0 | 212.4 | 529.2 |
| **Uganda**<br>*DP x 6* | VSL Y* | 3.6 | 3.7 | 4.2 | 96.3 | 169.9 | 443.3 |
|  | VSL SA1 | 2.1 | 2.3 | 2.8 | 94.8 | 168.5 | 441.8 |
|  | VSL SA2 | 5.9 | 6.0 | 6.5 | 98.6 | 172.3 | 445.6 |
|  | VSL SA3 | 9.2 | 9.3 | 9.8 | 101.9 | 175.5 | 448.9 |

### 6.2 Health benefit estimates

Our model estimated that implementing malaria chemoprevention in SAC aggregated across the ten HBHI countries for one year would avert an estimated 9.1 million malaria cases, 20,367 deaths, and 389,765 DALYs. The intervention would generate $245.1 million in morbidity-related savings and $5.53 billion in mortality risk protection benefits. The modeled number of malaria cases, deaths, and DALYs averted ranged from 256,515 to 2,995,407 cases, 443 to 8,119 deaths, and 8,496 to 155,341 DALYs, with the lowest and highest estimates from Mali and Nigeria, respectively (Table 1, Supplementary Table S6). Mortality risk reduction benefits would consistently outweigh morbidity-related benefits by at least tenfold in all countries, with the ratio approaching 30:1 in Tanzania, Nigeria, and Niger.

### 6.3 Educational gains

In the ten HBHI countries, our model found that malaria chemoprevention in SAC reduced absenteeism, resulting in an estimated 126,852 additional years of schooling (equivalent to 46.3 million additional school days) and $86.7 million in projected human capital gains (Table 1).

Based on our modeling of gains in foundational skills of literacy, malaria chemoprevention in SAC was estimated to generate 18.5 million LAYS in the aggregated ten HBHI countries, corresponding to $97 billion in human capital gains (Supplementary Table S6). The model projected that for every $100 invested, malaria chemoprevention in SAC could yield 5.03 LAYS. Among 150 education interventions analyzed globally, only four were estimated to deliver more LAYS per $100, one of which is school-based deworming. Programs that deliver three additional LAYS per $100 per child are considered highly cost-effective (Angrist et al. 2020).

### 6.4 Benefit-cost ratio and Net benefits

From a societal perspective, aggregated across the ten HBHI countries and using the absenteeism-based approach, our model estimated that implementing malaria chemoprevention in SAC would generate $5.7 billion in net benefits, resulting in a BCR of 14.28, classified as “good” according to the traffic light rating system. Two countries were predicted to have BCRs in the “excellent” category, equivalent to the top 20th percentile of interventions evaluated by the Copenhagen Consensus. Five countries fell into the “good” category and three into the “fair” category. Modelled country-level societal BCRs ranged from 3.71 in Uganda to 35.70 in Nigeria. The lowest BCR in Uganda’s DP scenario reflected the high costs of delivering DP six times annually, while Nigeria’s substantially higher BCR was driven by a combination of: (1) a high VSL due to higher GNI per capita; (2) a higher malaria case fatality rate (CFR); and (3) a higher malaria incidence. Across all countries, mortality risk reduction benefits accounted for most modeled benefits. The importance of drug choice is further explored in sensitivity analysis found in the Supplementary materials (Table S7).

### 6.5 Structural Sensitivity Analysis

Structural sensitivity analysis revealed that our modeled BCR estimates were highly sensitive to the method used to quantify educational gains. BCRs based on foundational skills in literacy improvements were up to 100 times higher than those based solely on reduced absenteeism. This result was not surprising, as literacy-based gains reflect improvements in test scores across the entire classroom cohort, assuming all children received a benefit from malaria chemoprevention in SAC. In contrast, absenteeism-based estimates capture only the individual-level benefit of additional school days for pupils who would otherwise have missed school due to malaria.

Discount rates had a significant impact on our models that estimated literacy-based BCRs, but had little effect on absenteeism-based estimates. The choice of value of a statistical life (VSL) method also influenced our models, in some cases doubling the BCRs, though this effect was modest compared to differences driven by the choice of educational outcome metric. Despite these structural variations, all modeled scenarios produced fair or good BCRs, with literacy-based BCRs consistently falling in the “excellent” range across all countries.

### 6.6 Probabilistic Sensitivity Analysis of Benefit-Cost Ratios

Performing a probabilistic sensitivity analysis with 10,000 iterations based on pooled input distributions, the simulated BCR for malaria chemoprevention in SAC across the ten HBHI countries was 11.00 (95% CI: 10.89–11.11), with a median of 9.94 and a range from 1.02 to 79.12. The relatively narrow CI for the “mean country” BCR reflects the design of this analysis: pooled inputs were used to simulate average conditions, and uncertainty was varied only around this profile. As a result, the CI represents the **precision of the simulated mean estimate**, rather than the full spread of possible outcomes. The distribution of BCRs was right-skewed (skewness = 1.44), with a kurtosis of 4.81, reflecting the presence of a small number of simulations with very high BCRs. The benefit-cost acceptability curve illustrates the probability that the intervention is cost-beneficial at varying BCR thresholds (Figure 4). While any BCR above 1 indicates that benefits outweigh costs, decision-makers often compare across competing interventions and may apply higher thresholds to prioritize those with the greatest relative returns. In our analysis, the acceptability curve indicated a >95% probability that malaria chemoprevention in SAC is cost-beneficial at a BCR threshold of 3, increasing to nearly 100% at a threshold of 15. The tornado diagram highlighted the case fatality rate, incidence rate, and intervention effect size as the most influential drivers of variation in the BCR, underscoring the importance of accurate parameter estimates and the potential to target programs based on these characteristics (Supplementary Figure 1).

**Figure 4.**
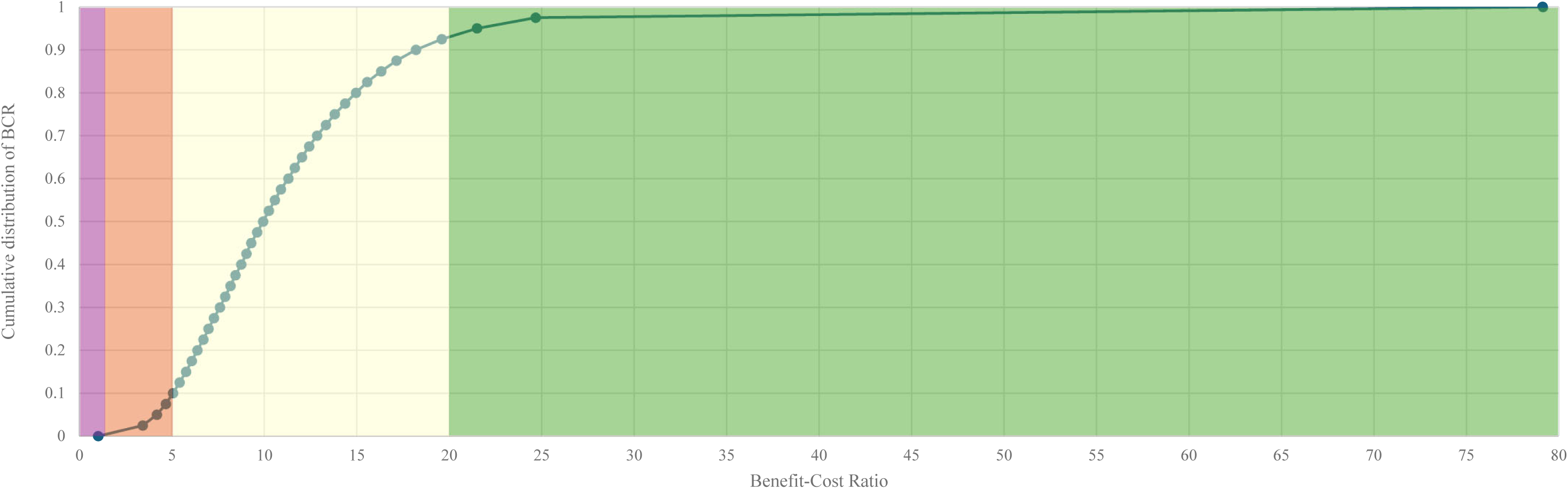
Benefit-cost acceptability curve using average input parameters across 10 high burden to high impact countries. The probabilistic sensitivity analysis was conducted on a simulated country profile to explore the uncertainty in model outputs based on average values or distributions derived from the 10 countries included in the analysis. Educational gains estimated using the absenteeism-approach. Green color: “excellent” rating; yellow color: “good” rating; orange: “fair” rating; purple: “poor” rating according to the Copenhagen Consensus ‘traffic light’ rating system.

### 6.7 Incremental cost-effectiveness ratios

From a provider perspective and using cost-effectiveness thresholds that range from 15-36% gross domestic product per capita (reflecting opportunity costs of health spending in LMICs), our models found that malaria chemoprevention in SAC in the ten HBHI countries had an ICER of $944 (range: $297-2,975) per DALY averted and was not cost-effective compared to no chemoprevention in any HBHI country or in the aggregated analysis (Supplementary Table S6) (Pichon-Riviere et al. 2023). However, these ICERs may underestimate the true value of the intervention, as they do not account for potential reductions in malaria transmission to other age groups. Cost per case averted in the ten HBHI countries ($40 per case averted; range: $26-67 per case averted), which is higher than cost per case averted for bednets ($7) and IPTi ($12), but lower than seasonal malaria chemoprevention ($145) (Conteh et al. 2021).

## 7 Conclusion

This modelled benefit-cost analysis of implementing malaria chemoprevention in SAC in ten HBHI countries demonstrated substantial economic value, with estimated societal net benefits of $5.7 billion and a BCR of 14.28. These benefits were primarily driven by averted mortality, with additional gains from improved educational outcomes due to reduced absenteeism. Probabilistic sensitivity analysis yielded a mean simulated generalized societal BCR of 11.00 (95% CI: 10.89–11.11). Although the “mean country” does not represent any single setting, it serves as a benchmark for typical HBHI conditions, complementing country-specific analyses by providing both a generalizable measure of robustness and context-specific estimates for policy. Our findings demonstrated how different approaches to valuing human capital gains from improved education led to varying rates of return, with the LAYS approach yielding BCRs up to 100 times higher, underscoring the importance of further empirical work and stronger data on learning gains to substantiate the LAYS framework. While malaria chemoprevention in SAC was cost-beneficial in all settings, its cost-effectiveness appeared more limited compared to interventions such as bednets. This study fills a critical evidence gap by presenting the first BCA on malaria chemoprevention in SAC and quantifying the broader impacts of malaria prevention. These findings provide policymakers and funders with a strong basis to prioritize chemoprevention targeting SAC as a high-return investment that advances both health and human capital in malaria-endemic regions.

## Data Availability

All data produced in the present study are available upon reasonable request to the authors.

## Acknowledgments

We acknowledge OpenPhilanthropy (GV673604850) for funding this work. We thank Michael Willoughby (RTI International) for his contributions to the development of Figure 1.

## Footnotes

1 We included the ten countries originally targeted by the WHO HBHI initiative in 2018, excluding India, which officially exited the initiative in 2024 following significant reductions in malaria burden. Although Sudan joined HBHI in 2022, we excluded it due to conflict-related disruptions since 2023. The ongoing conflict has also compromised data availability and reliability, limiting the representativeness of recent data.

## References

Angrist, Noam, Elisabetta Aurino, Harry A. Patrinos, George Psacharopoulos, Emiliana Vegas, Ralph Nordjo, and Brad Wong. 2023. “Improving Learning in Low- and Lower-Middle-Income Countries.” Journal of Benefit-Cost Analysis 14 (August):55–80. 10.1017/bca.2023.26.

Angrist, Noam, David K Evans, Deon Filmer, Rachel Glennerster, F Halsey, and Rogers Shwetlena Sabarwal. 2020. “How to Improve Education Outcomes Most Efficiently? A Comparison of 150 Interventions Using the New Learning-Adjusted Years of Schooling Metric.” http://www.worldbank.org/prwp.

Angrist, Noam, Matthew C. H. Jukes, Sian Clarke, R. Matthew Chico, Charles Opondo, Donald Bundy, and Lauren M. Cohee. 2023. “School-Based Malaria Chemoprevention as a Cost-Effective Approach to Improve Cognitive and Educational Outcomes: A Meta-Analysis.” In Draft, March. http://arxiv.org/abs/2303.10684.

Ayieko, Philip, Angela O Akumu, Ulla K Griffiths, and Mike English. 2009. “The Economic Burden of Inpatient Paediatric Care: Household and Provider Costs for Treatment of Pneumonia, Malaria and Meningitis.” Cost Effectiveness and Resource Allocation 7 (1): 3. 10.1186/1478-7547-7-3.

Baird, Matthew D., and John F. Pane. 2019. “Translating Standardized Effects of Education Programs Into More Interpretable Metrics.” Educational Researcher 48 (4): 217–28. 10.3102/0013189X19848729.

Barofsky, Jeremy, Tobenna D. Anekwe, and Claire Chase. 2020. “Malaria Eradication and Economic Outcomes in Sub-Saharan Africa: Evidence from Uganda.” Journal of Health Economics 44 (December):118–36. 10.1016/j.jhealeco.2015.08.002.

Bhatt, S, D J Weiss, E Cameron, D Bisanzio, B Mappin, and U Dalrymple. 2016. “Europe PMC Funders Group The Effect of Malaria Control on Plasmodium Falciparum in Africa between 2000 and 2015” 526 (7572): 207–11. 10.1038/nature15535.The.

Brooker, Simon J, Sian Clarke, Deepika Fernando, Caroline W Gitonga, Joaniter Nankabirwa, David Schellenberg, and Brian Greenwood. 2017. “Malaria in Middle Childhood and Adolescence.” In Disease Control Priorities, edited by Donald Bundy, Nilanthi de Silva, Susan Horton, Dean T Jamison, and George C Patton, Third. Washington, DC: World Bank. http://www.healthdata.org/sites/default/fi.

Bundy, Donald A. P., Nilanthi de Silva, Susan Horton, Dean T. Jamison, and George C. Patton. 2017. Disease Control Priorities, Third Edition (Volume 8): Child and Adolescent Health and Development. Edited by Donald A. P. Bundy, Nilanthi de Silva, Susan Horton, Dean T. Jamison, and George C. Patton. Washington, DC: World Bank. 10.1596/978-1-4648-0423-6.

Burkybile, Frank. 2025. “Malaria Is Poised for a Devastating Comeback in Africa’s ‘Worst Moment in 25 Years.’” BMJ 389 (April):r783. 10.1136/bmj.r783.

Caesar delos Trinos, John Paul, Dinh Ng-Nguyen, Luc E. Coffeng, Clare E.F. Dyer, Naomi Clarke, Rebecca Traub, Kate Halton, Virginia Wiseman, Caroline Watts, and Susana Vaz Nery. 2023. “Cost and Cost-Effectiveness Analysis of Mass Drug Administration Compared to School-Based Targeted Preventive Chemotherapy for Hookworm Control in Dak Lak Province, Vietnam.” The Lancet Regional Health - Western Pacific 41 (December):100913. 10.1016/j.lanwpc.2023.100913.

Chima, Reginald Ikechukwu, Catherine A Goodman, and Anne Mills. 2003. “The Economic Impact of Malaria in Africa: A Critical Review of the Evidence.” Health Policy 63 (1): 17–36. 10.1016/S0168-8510(02)00036-2.

Clarke, Siân E, Matthew C H Jukes, Kiambo Njagi, Lincoln Khasakhala, Bonnie Cundill, Julius Otido, Christopher Crudder, Benson B A Estambale, and Simon Brooker. 2008. “Eff Ect of Intermittent Preventive Treatment of Malaria on Health and Education in Schoolchildren: A Cluster-Randomised, Double-Blind, Placebo-Controlled Trial.” Www.Thelancet.Com. Vol. 372. www.thelancet.com.

Cohee, Lauren M., Charles Opondo, Siân E. Clarke, Katherine E. Halliday, Jorge Cano, Andrea G. Shipper, Breanna Barger-Kamate, et al. 2020. “Preventive Malaria Treatment among School-Aged Children in Sub-Saharan Africa: A Systematic Review and Meta-Analyses.” The Lancet Global Health 8 (12): e1499–1511. 10.1016/S2214-109X(20)30325-9.

Conteh, Lesong, Kathryn Shuford, Efundem Agboraw, Mara Kont, Jan Kolaczinski, and Edith Patouillard. 2021. “Costs and Cost-Effectiveness of Malaria Control Interventions: A Systematic Literature Review.” Value in Health 0 (0). 10.1016/j.jval.2021.01.013.

Copenhagen Consensus Center. (2021). Identifying Best Buys for Africa. https://copenhagenconsensus.com/sites/default/files/identifying_best_buys_-_copenhagen_consensus_center_2021.pdf

Evans, David K, and Fei Yuan. 2019. “Equivalent Years of Schooling A Metric to Communicate Learning Gains in Concrete Terms.” www.worldbank.org/research.

Ezennia, Ifeoma Jovita, Sunday Odunke Nduka, and Obinna Ikechukwu Ekwunife. 2017. “Cost Benefit Analysis of Malaria Rapid Diagnostic Test: The Perspective of Nigerian Community Pharmacists.” Malaria Journal 16 (1): 7. 10.1186/s12936-016-1648-0.

Fernando, Deepika, Damani de Silva, Richard Carter, Kamini N Mendis, and Rajitha Wickremasinghe. 2006. “A Randomized, Double-Blind, Placebo-Controlled, Clinical Trial of the Impact of Malaria Prevention on the Educational Attainment of School Children.” The American Journal of Tropical Medicine and Hygiene 74 (3): 386–93.

Fink, Günther, Evan Peet, Goodarz Danaei, Kathryn Andrews, Dana Charles McCoy, Christopher R. Sudfeld, Mary C. Smith Fawzi, Majid Ezzati, and Wafaie W. Fawzi. 2016. “Schooling and Wage Income Losses Due to Early-Childhood Growth Faltering in Developing Countries: National, Regional, and Global Estimates.” American Journal of Clinical Nutrition 104 (1): 104–12. 10.3945/ajcn.115.123968.

Galactionova, Katya, Thomas A. Smith, Don de Savigny, and Melissa A. Penny. 2017. “State of Inequality in Malaria Intervention Coverage in Sub-Saharan African Countries.” BMC Medicine 15 (1): 185. 10.1186/s12916-017-0948-8.

Gething, Peter W, Anand P Patil, David L Smith, Carlos A Guerra, Iqbal RF Elyazar, Geoffrey L Johnston, Andrew J Tatem, and Simon I Hay. 2011. “A New World Malaria Map: Plasmodium Falciparum Endemicity in 2010.” Malaria Journal 10 (1): 378. 10.1186/1475-2875-10-378.

GiveWell. 2021. “Evidence Actions Deworm the World Initiative - November 2021.” https://www.givewell.org/charities/deworm-world-initiative/November-2021-version#sources1155.

Global Fund. 2024a. “Indicative Reference Costs for Budgeting Purposes: International Freight, Insurance, and Quality Assurance.” https://www.theglobalfund.org/media/8985/ppm_freightinsurancequalityreferencecosts_list_en.pdf.

Global Fund. 2024b. “Pooled Procurement Mechanism Reference Pricing: Antimalarial Medicines.” https://www.theglobalfund.org/media/5812/ppm_actreferencepricing_table_en.pdf.

Gonahasa, Samuel, Martha Nassali, Catherine Maiteki-Sebuguzi, Jane F. Namuganga, Jimmy Opigo, Isaiah Nabende, Jaffer Okiring, et al. 2024. “LLIN Evaluation in Uganda Project (LLINEUP2): Association between Housing Construction and Malaria Burden in 32 Districts.” Malaria Journal 23 (1). 10.1186/s12936-024-05012-y.

Haacker, M., Hallett, T. B., & Atun, R. (2019). On discount rates for economic evaluations in global health. Health Policy and Planning. 10.1093/heapol/czz127

Halliday, Katherine E, Stefan S Witek-McManus, Charles Opondo, Austin Mtali, Elizabeth Allen, Andrew Bauleni, Saidi Ndau, et al. 2020. “Impact of School-Based Malaria Case Management on School Attendance, Health and Education Outcomes: A Cluster Randomised Trial in Southern Malawi.” BMJ Global Health 5 (1): e001666. 10.1136/bmjgh-2019-001666.

Hansen, Kristian Schultz, and Shunmay Yeung. 2009. “ACT Consortium Guidance on Collecting Household Costs.,” 1–11. http://www.actconsortium.org/data/files/household_costs.pdf.

Hodgson, Thomas A., and Mark R. Meiners. 1982. “Cost-of-Illness Methodology: A Guide to Current Practices and Procedures.”

Jamison, Dean T., Prabhat Jha, Ramanan Laxminarayan, and Toby Ord. 2013. “Infectious Disease, Injury, and Reproductive Health.” In Global Problems, Smart Solutions: Costs and Benefits, edited by Bjorn Lomborg. Cambridge Uniiversity Press.

Jo, Changik. 2014. “Cost-of-Illness Studies: Concepts, Scopes, and Methods.” Clinical and Molecular Hepatology 20 (4): 327–37. 10.3350/cmh.2014.20.4.327.

Korenromp, Eline, Avenir Health, Geneva Switzerland, Brad Wong, Nancy Dubosse, and Adamson S Muula. 2021. “The Cost-Benefit Analysis of Malaria Control Strategies in Malawi: A Scenario Comparison Using the Spectrum-Malaria Impact Modelling Tool.” Liverpool School of Tropical Medicine. Malawi: Prof Don Pascal Mathanga. www.afidep.org.

Kuecken, Maria, Josselin Thuilliez, and Marie Anne Valfort. 2021. “Disease and Human Capital Accumulation: Evidence from the Roll Back Malaria Partnership in Africa.” Economic Journal 131 (637): 2171–2202. 10.1093/ej/ueaa134.

Larg, Allison, and John R. Moss. 2011. “Cost-of-Illness Studies: A Guide to Critical Evaluation.” PharmacoEconomics 29 (8): 653–71. 10.2165/11588380-000000000-00000.

Lubell, Yoel, Hugh Reyburn, Hilda Mbakilwa, Rose Mwangi, Semkini Chonya, Christopher J M Whitty, and Anne Mills. 2008. “The Impact of Response to the Results of Diagnostic Tests for Malaria: Cost-Benefit Analysis.” BMJ 336 (7637): 202–5. 10.1136/bmj.39395.696065.47.

Lucas, Adrienne M. 2010. “Malaria Eradication and Educational Attainment: Evidence from Paraguay and Sri Lanka.” American Economic Journal: Applied Economics 2 (2): 46–71. 10.1257/app.2.2.46.

Maitra, Chandana, Andrew Hodge, and Eliana Jimenez Soto. 2016. “A Scoping Review of Cost Benefit Analysis in Reproductive, Maternal, Newborn and Child Health: What We Know and What Are the Gaps?” Health Policy and Planning 31 (10): 1530–47. 10.1093/heapol/czw078.

Makenga, Geofrey, Vito Baraka, Filbert Francis, Swabra Nakato, Samwel Gesase, George Mtove, Rashid Madebe, et al. 2023. “Effectiveness and Safety of Intermittent Preventive Treatment with Dihydroartemisinin–Piperaquine or Artesunate–Amodiaquine for Reducing Malaria and Related Morbidities in Schoolchildren in Tanzania: A Randomised Controlled Trial.” The Lancet Global Health 11 (8): e1277–89. 10.1016/S2214-109X(23)00204-8.

Mincer, Jacob A. 1974. “Schooling and Earnings.” In Schooling, Experience, and Earnings. National Bureau of Economic Research. http://www.nber.org/chapters/c1765?

Montresor, A., A.F. Gabrielli, A. Diarra, and D. Engels. 2010. “Estimation of the Cost of Large-Scale School Deworming Programmes with Benzimidazoles.” Transactions of the Royal Society of Tropical Medicine and Hygiene 104 (2): 129–32. 10.1016/j.trstmh.2009.10.007.

Morlino, Colette, Isabel Byrne, Jane Achan, Vito Baraka, Aissata Barry, Teun Bousema, Alioune Camara, et al. 2025. “Barriers to Uptake and Implementation of Malaria Chemoprevention in School-Aged Children: A Stakeholder Engagement Meeting Report.” Frontiers in Tropical Diseases 6:1480907. 10.3389/FITD.2025.1480907.

Mouzin, Eric, Richard Sedlmayr, John Miller, Rick Steketee, Paul Banda, Gilbert Chiyota, and Chuma Kabaghe. 2011. “Business Investing in Malaria Control: Economic Returns and a Healthy Workforce for Africa.”

Namuganga, Jane F., Joaniter I. Nankabirwa, Catherine Maiteki-Ssebuguzi, Samuel Gonahasa, Jimmy Opigo, Sarah G. Staedke, Damian Rutazaana, et al. 2022. “East Africa International Center of Excellence for Malaria Research: Impact on Malaria Policy in Uganda.” The American Journal of Tropical Medicine and Hygiene 107 (4_Suppl): 33–39. 10.4269/ajtmh.21-1305.

Nankabirwa, Joaniter, Simon J. Brooker, Sian E. Clarke, Deepika Fernando, Caroline W. Gitonga, David Schellenberg, and Brian Greenwood. 2014. “Malaria in SchoolLage Children in Africa: An Increasingly Important Challenge.” Tropical Medicine & International Health 19 (11): 1294–1309. 10.1111/tmi.12374.

National Institute for Medical Research, Tanzania. 2023. “The Use of Intermittent Preventive Treatment for Reducing Malaria Burden in School-Aged Children in Moderate and High Endemic Areas in Tanzania.” chrome-extension://efaidnbmnnnibpcajpcglclefindmkaj/https://nimr.or.tz/wp-content/uploads/2024/11/IPTsc-MALARIA-with-PG-inputs.pdf.

Patrinos, Harry Anthony, and George Psacharopoulos. 2020. “Returns to Education in Developing Countries.” In The Economics of Education, 53–64. Elsevier. 10.1016/B978-0-12-815391-8.00004-5.

Pichon-Riviere, Andres, Michael Drummond, Alfredo Palacios, Sebastián Garcia-Marti, and Federico Augustovski. 2023. “Determining the Efficiency Path to Universal Health Coverage: Cost-Effectiveness Thresholds for 174 Countries Based on Growth in Life Expectancy and Health Expenditures.” The Lancet Global Health 11 (6): e833–42. 10.1016/S2214-109X(23)00162-6.

Pradhan, Elina, and Dean T. Jamison. 2019. “Standardized Sensitivity Analysis in BCA: An Education Case Study.” Journal of Benefit-Cost Analysis 10 (March):206–23. 10.1017/bca.2019.5.

Pradhan, Elina, Elina M. Suzuki, Sebastián Martínez, Marco Schäferhoff, and Dean T. Jamison. 2017. “The Effects of Education Quantity and Quality on Child and Adult Mortality: Their Magnitude and Their Value.” In Disease Control Priorities, Third Edition (Volume 8): Child and Adolescent Health and Development, 423–40. The World Bank. 10.1596/978-1-4648-0423-6_ch30.

Purdy, Mark, Matthew Robinson, Kuangyi Wei, and David Rublin. 2013. “The Economic Case for Combating Malaria.” The American Society of Tropical Medicine and Hygiene 89 (5): 819–23. 10.4269/ajtmh.12-0689.

Rajkumar, Andrew Sunil, Christopher Guakler, and Jessica Tilahun. 2012. Combating Malnutrition in Ethiopia: An Evidence-Based Approach for Sustained Results. Washington, D.C.: World Bank.

Robinson, Lisa A, James K Hammitt, Michele Cecchini, Kalipso Chalkidou, Karl Claxton, Maureen Cropper, Patrick Hoang-Vu Eozenou, et al. 2019. “Reference Case Guidelines for Benefit-Cost Analysis in Global Health and Development.” https://sites.sph.harvard.edu/bcaguidelines/methods-and-cases/.

Setiawan, E., Cassidy-Seyoum, S. A., Thriemer, K., Carvalho, N., & Devine, A. (2024). A Systematic Review of Methods for Estimating Productivity Losses due to Illness or Caregiving in Low- and Middle-Income Countries. PharmacoEconomics, 42(8), 865–877. 10.1007/s40273-024-01402-x

Shretta, Rima, and Randolph Ngwafor Anye. 2023. “An Investment Case for the Scale-up and Use of Insecticide-Treated Nets Halfway into the SDG Targets.” Journal of Benefit-Cost Analysis 14 (December):16–54. 10.1017/bca.2023.23.

Sicuri, Elisa, Ana Vieta, Leandro Lindner, and Christophe Sauboin. 2011. “Economic Costs of Malaria in Children in Three Sub-Saharancountries: Ghana, Tanzania and Kenya.” Tropical Medicine and International Health 16 (SUPPL. 1): 117. http://ovidsp.ovid.com/ovidweb.cgi?T=JS&PAGE=reference&D=emed13&NEWS=N&AN=70589204.

Sixpence, Alick, Maclean Vokhiwa, Wangisani Kumalakwaanthu, Nicola J. Pitchford, Karl B. Seydel, Laurence S. Magder, Miriam K. Laufer, Don P. Mathanga, and Lauren M. Cohee. 2024. “Comparing Approaches for Chemoprevention for School-Based Malaria Control in Malawi: An Open Label, Randomized, Controlled Clinical Trial.” EClinicalMedicine 76 (October). 10.1016/j.eclinm.2024.102832.

Snyman, Katherine, Catherine Pitt, Angelo Aturia, Joyce Aber, Samuel Gonahasa, Jane Frances Namuganga, Joaniter Nankabirwa, et al. 2025. “Who Pays to Treat Malaria and How Much? Analysis of the Cost of Illness, Equity and Economic Burden of Malaria in Uganda.” Health Policy and Planning 40 (1): 52–65. 10.1093/heapol/czae093.

The Global Fund. 2023. “Pooled Procurement Mechanism Reference Pricing□: RDTs.” https://www.theglobalfund.org/media/7564/psm_hivrdtreferencepricing_table_en.pdf.

Tusting, Lucy S., Barbara Willey, Henry Lucas, John Thompson, Hmooda T. Kafy, Richard Smith, and Steve W. Lindsay. 2013. “Socioeconomic Development as an Intervention against Malaria: A Systematic Review and Meta-Analysis.” The Lancet 382 (9896): 963–72. 10.1016/S0140-6736(13)60851-X.

UNESCO Institute for Statistics. 2020. “Global Education Monitoring Report 2020: Inclusion and Education - All Means All.” https://www.unesco.org/gem-report/en/inclusion.

“UNESCO Institute for Statistics (UIS).” 2024. https://databrowser.uis.unesco.org/about.

United Nations, Department of Economic and Social Affairs, Population Division. 2024. “World Population Prospects 2024, Online Addition.” 2024. https://population.un.org/wpp/downloads?folder=Standard%20Projections&group=Population.

White, Michael T., Lesong Conteh, Richard Cibulskis, and Azra C. Ghani. 2011. “Costs and Cost-Effectiveness of Malaria Control Interventions - A Systematic Review.” Malaria Journal 10:1–14. 10.1186/1475-2875-10-337.

Wilkinson, Thomas, Mark J. Sculpher, Karl Claxton, Paul Revill, Andrew Briggs, John A. Cairns, Yot Teerawattananon, et al. 2016. “The International Decision Support Initiative Reference Case for Economic Evaluation: An Aid to Thought.” Value in Health 19 (8): 921–28. 10.1016/j.jval.2016.04.015.

World Bank. 2020. “COST-EFFECTIVE APPROACHES TO IMPROVE GLOBAL LEARNING What Does Recent Evidence Tell Us Are ‘Smart Buys’ for Improving Learning in Low-and Middle-Income Countries? Recommendations of the Global Education Evidence Advisory Panel.”

World Bank. 2025. “World Development Indicators.” 2025. https://databank.worldbank.org/source/world-development-indicators.

World Health Organization. 2011. “WHO-CHOICE Estimates of Cost for Inpatient and Outpatient Health Service Delivery.” https://www.who.int/choice/cost-effectiveness/inputs/health_service/en/.

World Health Organization. 2018. “High Burden to High Impact: A Targeted Malaria Response.” https://www.who.int/publications/i/item/WHO-CDS-GMP-2018.25.

World Health Organization. 2020. “Global Health Estimates 2019: DALY Methods.” https://www.who.int/docs/default-source/gho-documents/global-health-estimates/ghe2019_daly-methods.pdf.

World Health Organization. 2023. “WHO Guidelines for Malaria.” Who. Geneva. https://www.who.int/publications/i/item/guidelines-for-malaria.

World Health Organization. 2024. World Malaria Report 2024. World Health Organization. https://www.who.int/teams/global-malaria-programme/reports/world-malaria-report-2024.

World Health Organization. 2025. “Country-Specific Life Tables.” Global Health Observatory. 2025. https://apps.who.int/gho/data/node.main.LIFECOUNTRY?lang=en.

Zongo, Issaka, Paul Milligan, Yves Daniel Compaore, A. Fabrice Some, Brian Greenwood, Joel Tarning, Philip J. Rosenthal, Colin Sutherland, Francois Nosten, and Jean-Bosco Ouedraogo. 2015. “Randomized Noninferiority Trial of Dihydroartemisinin-Piperaquine Compared with Sulfadoxine-Pyrimethamine plus Amodiaquine for Seasonal Malaria Chemoprevention in Burkina Faso.” Antimicrobial Agents and Chemotherapy 59 (8): 4387–96. 10.1128/AAC.04923-14.

Zulaika, Garazi, Elizabeth Nyothach, Anna Maria van Eijk, Duolao Wang, Valarie Opollo, David Obor, Linda Mason, et al. 2023. “Menstrual Cups and Cash Transfer to Reduce Sexual and Reproductive Harm and School Dropout in Adolescent Schoolgirls in Western Kenya: A Cluster Randomised Controlled Trial.” EClinicalMedicine 65 (November):102261. 10.1016/j.eclinm.2023.102261.

