## Supplementary Materials for "A Smart Investment: The Health, Education, and Economic Returns of Malaria Chemoprevention in School-Aged Children Across Ten High-Burden Countries"

### Literature Review

#### Table S1 Benefit-cost ratios for malaria control interventions

ITN: Insecticide-treated bed net; PBO: piperonyl butoxide; IRS: Indoor residual spraying; ACT: artemisinin-based combination therapy

| **Source (author, year)** | **Modelled Years** | **Country** | **Intervention** | **BCR** |
| --- | --- | --- | --- | --- |
| Shretta et al., 2023 | 2023-2030 | 29 Highest burden countries in Africa | Scaling up ITNs by 10% | 48 |
| Korenromp et al., 2021 | 2021-2030 | Malawi | 100% PBO ITN distribution | 6 |
|  |  |  | Scaling up IRS in select districts | 1.7 |
|  |  |  | Mass media for improved care seeking for fever | 7.4 |
|  |  |  | Mass media for improved care seeking for fever + 100% PBO ITN distribution | 6.6 |
|  |  |  | Increasing frequency of mass ITN campaigns from 3-yearly to 2-yearly | <1 |
| Ezennia et al., 2017 | 2016 | Nigeria | Test-based malaria treatment | 6.7 |
| Purdy et al., 2013 | 2013-2035 | WHO regions | Roll Back Malaria (RBM) Global Malaria Action Plan (GMAP) to eradicate malaria | 6.1 |
| Rajkumar et al., 2012 | 2012 | Ethiopia | Bed Nets | 25.6 |
| Jamison et al., 2012 | 2012 | Low income countries, primarily Sub-Saharan Africa and South Asia | Subsidy on antimalarial drug: ACT | 35 |
| Mouzin et al., 2011 | 2001-2009 | Zambia (sugar plantation and copper mine) | Malaria prevention and control (including IRS and ITNs) | 1.04 |

### Parameter Tables

#### Table S2 Parameters for individual country analysis

For rows that have only one value, the same input was used for all countries. DRC: Democratic Republic of the Congo; * Incurred by government in public facilities, incurred by households in private facilities

|  | **Burkina Faso** | **Cameroon** | **DRC** | **Ghana** | **Mali** | **Mozambique** | **Niger** | **Nigeria** | **Tanzania** | **Uganda** | **Justification and (Source)** |
| --- | --- | --- | --- | --- | --- | --- | --- | --- | --- | --- | --- |
| **Model Assumptions** |  |  |  |  |  |  |  |  |  |  |  |
| Discount Rate | 5% |  |  |  |  |  |  |  |  |  |  |
| Primary school age range (years) | 6-11 | 6-11 | 6-11 | 6-11 | 7-12 | 6-12 | 7-12 | 6-11 | 7-13 | 6 - 12 | (“UNESCO Institute for Statistics (UIS)” 2024) |
| Total Population (2024) | 23,548,781 | 29,123,744 | 109,276,265 | 34,427,414 | 24,478,596 | 34,631,766 | 27,032,413 | 232,679,478 | 68,656,000 | 50,015,093 | (United Nations 2024) |
| % of population in primary school age range | 16.9% | 16.1% | 17.3% | 14.4% | 17.3% | 19.7% | 17.4% | 16.2% | 18.2% | 19.3% | (United Nations 2024) |
| Population cohort | 2,027,223 | 3,833,157 | 19,157,163 | 3,523,202 | 1,960,834 | 7,720,985 | 2,944,987 | 25,889,318 | 11,090,964 | 7,979,713 | Most recent data for number of primary school pupils multiplied by proportion enrolled in government funded school, adjusted for population growth using the SSA pupil growth rate for 2023; (“UNESCO Institute for Statistics (UIS)” 2024) |
| Number of primary schools distributing chemoprevention | 3,605 | 14,000 | 34,065 | 9,080 | 3,487 | 12,522 | 3,175 | 58,615 | 17,463 | 12,205 | Cameroon: Report only says ther are at least 14,000 public schools, not the exact number (Marist Foundation for International Solidarity 2023)  Ghana: Assume 49% are public (Ghana Investment Promotion Centre 2022)  Mozambique: Most recent data from 2017 (CEIC Data 2024)  Nigeria: Public primary schools only 2023 (Trustees of Columbia University 2025)  Tanzania: Enrollment in government primary schools(The United Republic of Tanzania Ministry of Education 2023)  Uganda: Government funded primary schools (Uganda Ministry of Education & Sports 2025)  Missing for Niger, Mali, DRC and Burkina Faso - so we used the mean pupil: school ratio from rest of countries and applied there. |
| Number of subnational units | 45 | 58 | 456 | 261 | 159 | 129 | 63 | 774 | 195 | 135 |  |
| **Health Outcomes** |  |  |  |  |  |  |  |  |  |  |  |
| **Malaria Incidence** |  |  |  |  |  |  |  |  |  |  |  |
| Incidence rate in SAC (case per 1000 child years) | 390 | 301 | 338 | 252 | 391 | 319 | 342 | 345 | 165 | 305 | Multiplied the total cases by the proportion of cases that are in SAC and divided by the proportion of population that is SAC, adjusted to 2024 levels using country-specific population growth rates; (World Health Organization 2024) |
| Rate ratio effect size of chemoprevention in SAC on clinical malaria | 0.665 | 0.665 | 0.665 | 0.665 | 0.665 | 0.665 | 0.665 | 0.665 | 0.8 | 0.5 | For DP six times annually: IRR: 0.5; 95% CI: 0.39-0.60(Cohee et al. 2020);  For DP three time annually: IRR: 0.8; 95% CI:0.71-0.91 (Makenga et al. 2023)  For SPAQ: we assume a proportionally lower protective effect compared to DP, reflecting its shorter duration of protection (4 weeks vs. 6 weeks)(Zongo et al. 2015). Accordingly, we apply two-thirds of the impact observed with DP |
| Proportion of cases of malaria that are in SAC age group | 0.187 | 0.187 | 0.187 | 0.187 | 0.195 | 0.228 | 0.195 | 0.187 | 0.234 | 0.228 | Enhanced malaria surveillance data from Uganda adjusted to the primary school age range in each country; (Namuganga et al. 2022) |
| **Disability Adjusted Life Years** | | |  |  |  |  |  |  |  |  |  |
| Percentage of cases that will become severe malaria in SAC | 2% |  |  |  |  |  |  |  |  |  | World malaria report estimated 1-3% of cases will become severe in models. Greenwood et al 1991 estimates 2% will become severe. (World Health Organization 2024) |
| Case fatality rate for uncomplicated malaria in SAC | 0.0020 | 0.0016 | 0.0020 | 0.0017 | 0.0017 | 0.0019 | 0.0044 | 0.0027 | 0.0030 | 0.0013 | Deaths / total number of cases (World Health Organization 2024) |
| Years of Life lost per death (discounted at 5%) | 18.70 | 18.57 | 18.63 | 18.76 | 18.70 | 18.26 | 18.70 | 18.82 | 18.82 | 18.76 | Standard methods from (Salomon et al. 2015; World Health Organization 2020) using data from country-specific WHO life tables. |
| Disability weight for a uncomplicated case of malaria | 0.053 |  |  |  |  |  |  |  |  |  | Global Burden of Disease Study (Salomon et al. 2015) |
| Disability weight for a severe case of malaria | 0.211 |  |  |  |  |  |  |  |  |  | Global Burden of Disease Study (Salomon et al. 2015) |
| Average length of uncomplicated malaria episode (years) | 0.014 |  |  |  |  |  |  |  |  |  | Global Burden of Disease Study (Salomon et al. 2015) |
| Average length of severe malaria episode (years) | 0.0281 |  |  |  |  |  |  |  |  |  | Global Burden of Disease Study (Salomon et al. 2015) |
| **Learning Outcomes** |  |  |  |  |  |  |  |  |  |  |  |
| Number of days lost school due to one episode of uncomplicated malaria | 5 |  |  |  |  |  |  |  |  |  | (Korenromp et al. 2021) |
| Number of days lost to school due to one episode of severe malaria | 10 |  |  |  |  |  |  |  |  |  | (Korenromp et al. 2021) |
| Expected years of school | 7 | 8.7 | 9.1 | 12.2 | 5.2 | 7.6 | 5.5 | 10.2 | 7.2 | 6.8 | (World Bank 2025a) |
| Learning adjustment | 0.8 |  |  |  |  |  |  |  |  |  | Benchmark of 0.8 standard deviations to represent high-performing learning rates (Evans and Yuan 2019; Angrist et al. 2020) |
| Estimated returns to schooling (% income for one more year) | 0.039 | 0.054 | 0.052 | 0.047 | 0.039 | 0.083 | 0.085 | 0.038 | 0.076 | 0.12 | Based on Mincerian estimates (Fink et al. 2016) |
| **Costs** |  |  |  |  |  |  |  |  |  |  |  |
| **Intervention Costs** |  |  |  |  |  |  |  |  |  |  |  |
| Cost per dose of chemoprevention drugs | $0.28 | $0.28 | $0.28 | $0.28 | $0.28 | $0.28 | $0.28 | $0.28 | $1.46 | $1.46 | (Global Fund 2024b) |
| Number of doses of chemoprevention drugs per student -annually | 6 | 6 | 6 | 6 | 6 | 6 | 6 | 6 | 3 | 6 | Assume six doses in all countries except for Tanzania, where three doses are currently administered |
| Intervention Coverage | 90% |  |  |  |  |  |  |  |  |  | Based on coverage in school based de-worming campaigns. (GiveWell 2021) |
| % Wastage of drugs | 5% |  |  |  |  |  |  |  |  |  | (Temperley et al. 2008) |
| Cost of freight and insurance (as % of medicines) | 10% |  |  |  |  |  |  |  |  |  | (Montresor et al. 2010; Global Fund 2024a) |
| Percentage of parents that attend community sensitization meeting | 50% |  |  |  |  |  |  |  |  |  | Authors assumption |
| Monthly Teacher salary | $687 | $322 | $50 | $117 | $232 | $322 | $430 | $203 | $551 | $212 | Burkina Faso, DRC, Ghana, Niger, Nigeria, Tanzania & Uganda: (Evans et al. 2020) Mali: (Oxfam International 2008) Cameroon & Mozambique: used mean from Evans et al. 2020 |
| Number of teacher-hours used to develiver chemoprevention per round per school | 12 |  |  |  |  |  |  |  |  |  | Assume six teacher per school spend 2 hours each round(Montresor et al. 2010) |
| Number of teachers per primary school to be trained | 12 |  |  |  |  |  |  |  |  |  | Authors assumption |
| **Morbidity-related cost-savings** | | | | |  |  |  |  |  |  |  |
| Probability of seeking care | 0.75 | 0.61 | 0.51 | 0.65 | 0.53 | 0.64 | 0.67 | 0.73 | 0.78 | 0.87 | Probability of seeking care for children under the age of 5 with a fever. (U.S. Agency for International Development 2025) 2022 DHS for Ghana, Mozambique, Tanzania 2022 MIS for Cameroon 2021 DHS for Burkina Faso 2021 MIS for Mali, Niger, Nigeria 2018 MIS for Uganda  2013/4 DHS for Democratic Republic of Congo |
| Percentage of care sought at publicly funded health centers | 0.94 | 0.31 | 0.49 | 0.39 | 0.42 | 0.94 | 0.81 | 0.37 | 0.63 | 0.43 | Percentage of cases that sought case at publicly funded health centers for children under 5 with a fever (U.S. Agency for International Development 2025) 2022 DHS for Ghana, Mozambique, Tanzania 2022 MIS for Cameroon 2021 DHS for Burkina Faso 2021 MIS for Mali, Niger, Nigeria 2018 MIS for Uganda  2013/4 DHS for Democratic Republic of Congo |
| Cost of RDT | $0.39 |  |  |  |  |  |  |  |  |  | pf/pan rapid diagnostics test (RDT) (The Global Fund 2023) |
| Cost of medicine for uncomplicated malaria * | $0.50 |  |  |  |  |  |  |  |  |  | All countries recommend one course of artemether-lumefantrine (AL). Used pricing midpoint; range 0.22 - 0.65.(Global Fund 2024b) |
| Cost of medicine for severe malaria* | $11.78 |  |  |  |  |  |  |  |  |  | All countries recommend artusenate (AS). Assumes 6 vials of AS; no range of prices for AS (Global Fund 2024b) |
| Cost per uncomplicated case consultation fee* | $2.38 | $3.41 | $1.80 | $17.42 | $2.54 | $2.72 | $1.31 | $10.09 | $4.13 | $3.94 | WHO-CHOICE (mean of bed and no bed) inflated to 2023 USD(World Health Organization 2011) |
| Cost of transportation and food for uncomplicated malaria (to households) | $0.50 |  |  |  |  |  |  |  |  |  | (Snyman et al. 2025) |
| Cost of transportation and food for severe malaria (to households) | $1.00 |  |  |  |  |  |  |  |  |  | (Snyman et al. 2025) |
| Cost of hospitalization per day in public facility (to government) | $9.47 | $16.10 | $4.79 | $75.49 | $9.96 | $9.63 | $4.14 | $49.08 | $16.91 | $15.58 | WHO-CHOICE; Cost of hospitalization per day in public facility (2010 PPP I$) Primary Hospital for primary hospital converted to 2023 USD (World Health Organization 2011) |
| Duration of hospital stay for severe malaria (days) | 4 |  |  |  |  |  |  |  |  |  | (White et al. 2011) |
| Value of day of lost productivity to households | $7.23 | $14.52 | $4.27 | $20.41 | $6.93 | $4.19 | $4.99 | $16.49 | $10.79 | $8.33 | GNI per capita / 365 days |
| Number of days of lost productivity for untreated malaria | 2 |  |  |  |  |  |  |  |  |  | Mid-point of values reported various papers (Ayieko et al. 2009; Chima, Goodman, and Mills 2003; Sicuri et al. 2011; Snyman et al. 2025) |
| Number of days of lost productivity for uncomplicated malaria | 2 |  |  |  |  |  |  |  |  |  | Mid-point of values reported various papers (Ayieko et al. 2009; Chima, Goodman, and Mills 2003; Sicuri et al. 2011; Snyman et al. 2025) |
| Number of days of caregiver productivity for severe malaria | 5 |  |  |  |  |  |  |  |  |  | (White et al. 2011) |
| **Mortality-related cost savings** | | | |  |  |  |  |  |  |  |  |
| Mean age of death | 9 |  |  |  |  |  |  |  |  |  | Median of primary school age range |
| Minimum age to start working | 15 |  |  |  |  |  |  |  |  |  | Assumption |
| GNI per capita | $2,640 | $5,300 | $1,560 | $7,450 | $2,530 | $1,530 | $1,820 | $6,020 | $3,940 | $3,040 | GNI per capita in current USD (World Bank 2025b) |
| Non health GNI | $2,846 | $2,921 | $2,925 | $2,914 | $2,904 | $2,765 | $2,863 | $2,916 | $2,938 | $2,898 | Atlas methods, in current USD (World Bank 2025b) |
| GNI per capita annual growth rate | 3.20% | 1.20% | 6.70% | 3.20% | 1.00% | 0.80% | 2.60% | 2.90% | 2.00% | 2.60% | Not available for Nigeria, so used the GDP growth rate (2023) (World Bank 2025b) |
| **Cost-effectiveness thresholds** | | | | |  |  |  |  |  |  |  |
| Cost-effectiveness threshold | $230.53 | $220.51 | $150.35 | $230.53 | $160.37 | $360.83 | $250.58 | $170.39 | $260.60 | $170.39 | Cost-effectiveness threshold per QALY in US dollars, as a proportion of GDP per capita converted to 2023 USD(Pichon-Riviere et al. 2023) |

#### Table S3 Derivation of Country-Specific VSL and Age-Specific VSL

Table S3 details the country-specific calculations used to extrapolate VSL from the U.S. reference value and to derive age-specific VSL estimates using country life tables. Extrapolated VSL calculated as VSL_US_ × (GNI_c_ / GNI_US_)^1.5, using VSL_US_ = 13,710,363 USD and GNI_US_ = 84,450 USD. VSLY calculated as VSL divided by undiscounted remaining life expectancy at average adult age

|  | **Burkina Faso** | **Cameroon** | **DRC** | **Ghana** | **Mali** | **Mozambique** | **Niger** | **Nigeria** | **Tanzania** | **Uganda** |
| --- | --- | --- | --- | --- | --- | --- | --- | --- | --- | --- |
| GNI | $2,640 | $5,300 | $1,560 | $7,450 | $2,530 | $1,530 | $1,820 | $6,020 | $3,940 | $3,040 |
| Elasticity | 1.5 | 1.5 | 1.5 | 1.5 | 1.5 | 1.5 | 1.5 | 1.5 | 1.5 | 1.5 |
| Extrapolated VSL | $75,780 | $215,558 | $34,422 | $359,239 | $71,094 | $33,434 | $43,377 | $260,942 | $138,164 | $93,640 |
| Adult remaining life expectancy | 34 | 32 | 33 | 35 | 34 | 28 | 34 | 36 | 36 | 35 |
| VSLY | $2,253 | $6,659 | $1,041 | $10,398 | $2,077 | $1,207 | $1,282 | $7,316 | $3,850 | $2,690 |
| Remaining life expectancy at age 9 | 64 | 62 | 63 | 65 | 64 | 58 | 64 | 66 | 66 | 65 |
| Age-specific VSL | $143,373 | $415,319 | $65,650 | $671,182 | $133,402 | $69,648 | $81,847 | $480,409 | $253,661 | $174,327 |

Extrapolated VSL calculated as VSL_US_ × (GNI_c_ / GNI_US_)^1.5, using VSL_US_ = 13,710,363 USD and GNI_US_ = 84,450 USD. VSLY calculated as VSL divided by undiscounted remaining life expectancy at average adult age

#### Table S4 Parameters and distributions for probabilistic sensitivity analysis

All values are on the natural scale. Lognormal distributions were parameterized using these values; SD refers to the natural-scale standard deviation unless otherwise noted. Triangular, Beta, and Gamma distributions use standard parameterizations. “N/A” indicates not applicable

| **Parameter** | **Mean Value** | **Distribution** | **Max** | **Min** | **Alpha** | **Beta** | **Theta** | **SD** | **Justification and Source** |
| --- | --- | --- | --- | --- | --- | --- | --- | --- | --- |
| Model Assumptions |  |  |  |  |  |  |  |  |  |
| Population cohort | 8,612,754 | N/A | N/A | N/A | N/A | N/A | N/A | N/A |  |
| Number of primary schools distributing chemoprevention | 16,822 | N/A | N/A | N/A | N/A | N/A | N/A | N/A |  |
| Number of subnational units | 228 | N/A | N/A | N/A | N/A | N/A | N/A | N/A |  |
| Health Outcomes |  |  |  |  |  |  |  |  |  |
| Incidence rate in SAC (case per 1000 child years) | 315 | Lognormal | N/A | N/A | N/A | N/A | N/A | 63.4082 |  |
| Rate ratio effect size of chemoprevention in SAC on clinical malaria | 0.66 | Lognormal | N/A | N/A | N/A | N/A | N/A | 0.06735 |  |
| Proportion of cases of malaria that are in SAC age group | 0.202 | N/A | N/A | N/A | N/A | N/A | N/A | N/A |  |
| Percentage of cases that will become severe malaria in SAC | 2% | Triangle | 1% | 3% | N/A | N/A | N/A | N/A | Min. and max. taken from range used in World Malaria Report (World Health Organization 2024) |
| Case fatality rate for uncomplicated malaria in SAC | 0.0022 | Beta | N/A | N/A | 5.86 | 2622.04 | N/A | N/A |  |
| **Learning Outcomes** |  |  |  |  |  |  |  |  |  |
| Number of days lost school due to one episode of uncomplicated malaria | 5 | Triangle | 2 | 6 | N/A | N/A | N/A | N/A | (Halliday et al. 2020) |
| Number of days lost to school due to one episode of severe malaria | 10 | Triangle | 7 | 12 | N/A | N/A | N/A | N/A | (Halliday et al. 2020) |
| Expected years of school | 8.0 | N/A | N/A | N/A | N/A | N/A | N/A | N/A |  |
| Estimated returns to schooling (% income for one more year) | 0.063 | Beta | N/A | N/A | 5.14 | 75.45 | N/A | 0.0251 |  |
| **Costs** |  |  |  |  |  |  |  |  |  |
| **Intervention Costs** |  |  |  |  |  |  |  |  |  |
| Cost per dose of chemoprevention drugs | $0.52 | N/A | N/A | N/A | N/A | N/A | N/A | N/A |  |
| Number of doses of IPTsc drugs per student annually | 5.7 | N/A | N/A | N/A | N/A | N/A | N/A | N/A |  |
| Intervention coverage | 90% | Triangle | 79% | 93% | N/A | N/A | N/A | N/A | Deworming campaigns report coverage levels that range from 79% (Pakistan 2019) to 93% (Kenya 2014-2019) (GiveWell 2021) |
| % Wastage of drugs | 5% | Triangle | 2% | 10% | N/A | N/A | N/A | N/A | Min. based on author’s assumptions; Max. from (Temperley et al. 2008) |
| Cost of freight and insurance (as % of medicines) | 10% | Triangle | 5% | 14% | N/A | N/A | N/A | N/A | Min. based on authors assumptions; Max. from median of essential medicines list(Global Fund 2024a) |
| Percentage of parents that attend community sensitization meeting | 50% | Triangle | 40% | 80% | N/A | N/A | N/A | N/A |  |
| Monthly Teacher salary | 313 | Gamma | N/A | N/A | 2.47 | N/A | 117.35 | 186.1178 |  |
| Number of teacher-hours used to deliver chemoprevention per round per school | 12 | Triangle | 6 | 18 | N/A | N/A | N/A | N/A |  |
| Number of teachers per primary school to be trained | 12 | Triangle | 6 | 18 | N/A | N/A | N/A | N/A |  |
| **Morbidity-related cost-savings** |  |  |  |  |  |  |  |  |  |
| Probability of seeking care | 0.67 | Beta | N/A | N/A | 10.6 | 5.14 | N/A | 0.10645 |  |
| Percentage of care sought at publicly funded health centers | 0.57 | Beta | N/A | N/A | 1.83 | 1.37 | N/A | 0.22893 |  |
| Cost per uncomplicated case consultation fee | $4.97 | Gamma | N/A | N/A | 0.982 | N/A | 5.065 | 4.759953 |  |
| Cost of transportation and food for uncomplicated malaria (to households) | $0.50 | Triangle | $0.25 | $1.00 | N/A | N/A | N/A | N/A |  |
| Cost of transportation and food for severe malaria (to households) | $1.00 | Triangle | $0.50 | $2.00 | N/A | N/A | N/A | N/A |  |
| Cost of hospitalization per day in public facility (to government) | $21.12 | Gamma | N/A | N/A | 0.834 | N/A | 25.04 | 21.81502 |  |
| Duration of hospital stay for severe malaria (days) | 4 | Triangle | 3 | 5 | N/A | N/A | N/A | N/A | Min. and max. from range found in (White et al. 2011) |
| Value of day of lost productivity to households | $9.82 | Gamma | N/A | N/A | 3.062 | N/A | 3.205 | 5.317935 | Base case: GNI per capita/365; Lower:50% GINI/365 |
| Number of days of caregiver lost productivity for untreated malaria | 2 | Triangle | 1 | 3 | N/A | N/A | N/A | N/A | Min. and max. from range found in various papers (Ayieko et al. 2009; Chima, Goodman, and Mills 2003; Sicuri et al. 2011; Snyman et al. 2025) |
| Number of days of caregiver lost productivity for uncomplicated malaria | 2 | Triangle | 1 | 3 | N/A | N/A | N/A | N/A | Min. and max. from range found in various papers (Ayieko et al. 2009; Chima, Goodman, and Mills 2003; Sicuri et al. 2011; Snyman et al. 2025) |
| Number of days of caregiver productivity for severe malaria | 5 | Triangle | 3 | 7 | N/A | N/A | N/A | N/A | Min. and max. from range found in (White et al. 2011) |
| **Mortality-related cost savings** |  |  |  |  |  |  |  |  |  |
| Mean age of death | 9 | N/A | N/A | N/A | N/A | N/A | N/A | N/A |  |
| Minimum age to start working | 15 | N/A | N/A | N/A | N/A | N/A | N/A | N/A |  |
| GNI per capita (2023) | $3,583 | N/A | N/A | N/A | N/A | N/A | N/A | N/A |  |
| GNI per capita annual growth rate (2023 value) | 3% | N/A | N/A | N/A | N/A | N/A | N/A | N/A |  |

***References***

Angrist, Noam, David K Evans, Deon Filmer, Rachel Glennerster, F Halsey, and Rogers Shwetlena Sabarwal. 2020. “How to Improve Education Outcomes Most Efficiently? A Comparison of 150 Interventions Using the New Learning-Adjusted Years of Schooling Metric.” http://www.worldbank.org/prwp.

Ayieko, Philip, Angela O Akumu, Ulla K Griffiths, and Mike English. 2009. “The Economic Burden of Inpatient Paediatric Care: Household and Provider Costs for Treatment of Pneumonia, Malaria and Meningitis.” *Cost Effectiveness and Resource Allocation* 7 (1): 3. https://doi.org/10.1186/1478-7547-7-3.

CEIC Data. 2024. “Mozambique Education Statistics.” 2024. https://www.ceicdata.com/en/mozambique/education-statistics/no-of-public-schools-primary-first-level.

Chima, Reginald Ikechukwu, Catherine A Goodman, and Anne Mills. 2003. “The Economic Impact of Malaria in Africa: A Critical Review of the Evidence.” *Health Policy* 63 (1): 17–36. https://doi.org/10.1016/S0168-8510(02)00036-2.

Cohee, Lauren M., Charles Opondo, Siân E. Clarke, Katherine E. Halliday, Jorge Cano, Andrea G. Shipper, Breanna Barger-Kamate, et al. 2020. “Preventive Malaria Treatment among School-Aged Children in Sub-Saharan Africa: A Systematic Review and Meta-Analyses.” *The Lancet Global Health* 8 (12): e1499–1511. https://doi.org/10.1016/S2214-109X(20)30325-9.

Evans, David K, and Fei Yuan. 2019. “Equivalent Years of Schooling A Metric to Communicate Learning Gains in Concrete Terms.” www.worldbank.org/research.

Evans, David K, Fei Yuan, Deon Filmer, World Bank, Lawrence Katz, Owen Ozier, Halsey Rogers, et al. 2020. “Are Teachers in Africa Poorly Paid? Evidence from 15 Countries.” www.cgdev.orgwww.cgdev.org.

Fink, Günther, Evan Peet, Goodarz Danaei, Kathryn Andrews, Dana Charles McCoy, Christopher R. Sudfeld, Mary C. Smith Fawzi, Majid Ezzati, and Wafaie W. Fawzi. 2016. “Schooling and Wage Income Losses Due to Early-Childhood Growth Faltering in Developing Countries: National, Regional, and Global Estimates.” *American Journal of Clinical Nutrition* 104 (1): 104–12. https://doi.org/10.3945/ajcn.115.123968.

Ghana Investment Promotion Centre. 2022. “Ghana’s Education Sector Report.” https://www.gipc.gov.gh/wp-content/uploads/2022/12/Ghanas-Education-Sector-Report.pdf.

GiveWell. 2021. “Evidence Actions Deworm the World Initiative - November 2021.” https://www.givewell.org/charities/deworm-world-initiative/November-2021-version#sources1155.

Global Fund. 2024a. “Indicative Reference Costs for Budgeting Purposes: International Freight, Insurance, and Quality Assurance.” https://www.theglobalfund.org/media/8985/ppm_freightinsurancequalityreferencecosts_list_en.pdf.

———. 2024b. “Pooled Procurement Mechanism Reference Pricing: Antimalarial Medicines.” https://www.theglobalfund.org/media/5812/ppm_actreferencepricing_table_en.pdf.

Halliday, Katherine E, Stefan S Witek-McManus, Charles Opondo, Austin Mtali, Elizabeth Allen, Andrew Bauleni, Saidi Ndau, et al. 2020. “Impact of School-Based Malaria Case Management on School Attendance, Health and Education Outcomes: A Cluster Randomised Trial in Southern Malawi.” *BMJ Global Health* 5 (1): e001666. https://doi.org/10.1136/bmjgh-2019-001666.

Korenromp, Eline, Avenir Health, Geneva Switzerland, Brad Wong, Nancy Dubosse, and Adamson S Muula. 2021. “The Cost-Benefit Analysis of Malaria Control Strategies in Malawi: A Scenario Comparison Using the Spectrum-Malaria Impact Modelling Tool.” *Liverpool School of Tropical Medicine*. Malawi: Prof Don Pascal Mathanga. www.afidep.org.

Makenga, Geofrey, Vito Baraka, Filbert Francis, Swabra Nakato, Samwel Gesase, George Mtove, Rashid Madebe, et al. 2023. “Effectiveness and Safety of Intermittent Preventive Treatment with Dihydroartemisinin–Piperaquine or Artesunate–Amodiaquine for Reducing Malaria and Related Morbidities in Schoolchildren in Tanzania: A Randomised Controlled Trial.” *The Lancet Global Health* 11 (8): e1277–89. https://doi.org/10.1016/S2214-109X(23)00204-8.

Marist Foundation for International Solidarity. 2023. “Universal Periodic Review (UPR) Cameroon Fourth Cycle 44th Session (6-17 Nov. 2023) Focus Equity of Access to Education and Increasing Quality of Education.” https://www.stopblablacam.com/society/0309-9293-cameroon-s-govt-made-a-lot-of-progress-in-improving-the-.

Montresor, A., A.F. Gabrielli, A. Diarra, and D. Engels. 2010. “Estimation of the Cost of Large-Scale School Deworming Programmes with Benzimidazoles.” *Transactions of the Royal Society of Tropical Medicine and Hygiene* 104 (2): 129–32. https://doi.org/10.1016/j.trstmh.2009.10.007.

Namuganga, Jane F., Joaniter I. Nankabirwa, Catherine Maiteki-Ssebuguzi, Samuel Gonahasa, Jimmy Opigo, Sarah G. Staedke, Damian Rutazaana, et al. 2022. “East Africa International Center of Excellence for Malaria Research: Impact on Malaria Policy in Uganda.” *The American Journal of Tropical Medicine and Hygiene* 107 (4_Suppl): 33–39. https://doi.org/10.4269/ajtmh.21-1305.

Oxfam International. 2008. “Delivering Education for All in Mali.” https://www-cdn.oxfam.org/s3fs-public/file_attachments/education-for-all-mali-report-0906_9.pdf.

Pichon-Riviere, Andres, Michael Drummond, Alfredo Palacios, Sebastián Garcia-Marti, and Federico Augustovski. 2023. “Determining the Efficiency Path to Universal Health Coverage: Cost-Effectiveness Thresholds for 174 Countries Based on Growth in Life Expectancy and Health Expenditures.” *The Lancet Global Health* 11 (6): e833–42. https://doi.org/10.1016/S2214-109X(23)00162-6.

Robinson, Lisa A, James K Hammitt, Michele Cecchini, Kalipso Chalkidou, Karl Claxton, Maureen Cropper, Patrick Hoang-Vu Eozenou, et al. 2019. “Reference Case Guidelines for Benefit-Cost Analysis in Global Health and Development.” https://sites.sph.harvard.edu/bcaguidelines/methods-and-cases/.

Salomon, Joshua A., Juanita A. Haagsma, Adrian Davis, Charline Maertens de Noordhout, Suzanne Polinder, Arie H. Havelaar, Alessandro Cassini, et al. 2015. “Disability Weights for the Global Burden of Disease 2013 Study.” *The Lancet Global Health* 3 (11): e712–23. https://doi.org/10.1016/S2214-109X(15)00069-8.

Sicuri, Elisa, Ana Vieta, Leandro Lindner, and Christophe Sauboin. 2011. “Economic Costs of Malaria in Children in Three Sub-Saharancountries: Ghana, Tanzania and Kenya.” *Tropical Medicine and International Health* 16 (SUPPL. 1): 117. http://ovidsp.ovid.com/ovidweb.cgi?T=JS&PAGE=reference&D=emed13&NEWS=N&AN=70589204.

Snyman, Katherine, Catherine Pitt, Angelo Aturia, Joyce Aber, Samuel Gonahasa, Jane Frances Namuganga, Joaniter Nankabirwa, et al. 2025. “Who Pays to Treat Malaria and How Much? Analysis of the Cost of Illness, Equity and Economic Burden of Malaria in Uganda.” *Health Policy and Planning* 40 (1): 52–65. https://doi.org/10.1093/heapol/czae093.

Temperley, Matilda, Dirk H Mueller, J Kiambo Njagi, Willis Akhwale, Siân E Clarke, Matthew CH Jukes, Benson BA Estambale, and Simon Brooker. 2008. “Costs and Cost-Effectiveness of Delivering Intermittent Preventive Treatment through Schools in Western Kenya.” *Malaria Journal* 7 (1): 196. https://doi.org/10.1186/1475-2875-7-196.

The Global Fund. 2023. “Pooled Procurement Mechanism Reference Pricing : RDTs.” https://www.theglobalfund.org/media/7564/psm_hivrdtreferencepricing_table_en.pdf.

The United Republic of Tanzania Ministry of Education, Science and Technology. 2023. “Basic Education Data 2023.” 2023. https://www.tamisemi.go.tz/singleministers/basic_education_data_2023.

Trustees of Columbia University. 2025. “GRID3 Data Portal.” https://data.grid3.org/datasets/8aef208577664f859c44a18f61238a90_0/explore?

Uganda Ministry of Education & Sports. 2025. “Uganda Educational Management Information System .” 2025. https://emis.go.ug/.

“UNESCO Institute for Statistics (UIS).” 2024. https://databrowser.uis.unesco.org/about.

United Nations, Department of Economic and Social Affairs, Population Division. 2024. “World Population Prospects 2024, Online Addition.” 2024. https://population.un.org/wpp/downloads?folder=Standard%20Projections&group=Population.

U.S. Agency for International Development. 2025. “The DHS Program STATcompiler.” 2025. http://www.statcompiler.com.

White, Michael T., Lesong Conteh, Richard Cibulskis, and Azra C. Ghani. 2011. “Costs and Cost-Effectiveness of Malaria Control Interventions - A Systematic Review.” *Malaria Journal* 10:1–14. https://doi.org/10.1186/1475-2875-10-337.

World Bank. 2025a. “Gender Data Portal.” 2025. https://genderdata.worldbank.org/en/indicator/hd-hci-eyrs?gender=total.

———. 2025b. “World Development Indicators.” 2025. https://databank.worldbank.org/source/world-development-indicators.

World Health Organization. 2011. “WHO-CHOICE Estimates of Cost for Inpatient and Outpatient Health Service Delivery.” https://www.who.int/choice/cost-effectiveness/inputs/health_service/en/.

———. 2020. “Global Health Estimates 2019: DALY Methods.” https://www.who.int/docs/default-source/gho-documents/global-health-estimates/ghe2019_daly-methods.pdf.

———. 2024. *World Malaria Report 2024*. World Health Organization. https://www.who.int/teams/global-malaria-programme/reports/world-malaria-report-2024.

Zongo, Issaka, Paul Milligan, Yves Daniel Compaore, A. Fabrice Some, Brian Greenwood, Joel Tarning, Philip J. Rosenthal, Colin Sutherland, Francois Nosten, and Jean-Bosco Ouedraogo. 2015. “Randomized Noninferiority Trial of Dihydroartemisinin-Piperaquine Compared with Sulfadoxine-Pyrimethamine plus Amodiaquine for Seasonal Malaria Chemoprevention in Burkina Faso.” *Antimicrobial Agents and Chemotherapy* 59 (8): 4387–96. https://doi.org/10.1128/AAC.04923-14.

### Results

#### Table S5 Annual intervention cost per pupil (societal perspective)

*All costs presented in 2023 USD.*

|  |  | Cost Category | | | | | | | Total cost | Cost per pupil | Cost per dose | Cost per dose delivered without drugs |
| --- | --- | --- | --- | --- | --- | --- | --- | --- | --- | --- | --- | --- |
| Country | Perspective | Drugs | Human resources | Monitoring and evaluation | Storage | Supplies and materials | Transportation | Community sensitization |  |  |  |  |
| Nigeria | Household | $0 | $0 | $0 | $0 | $0 | $0 | $26,687,276 | $26,687,276 | $1.03 | $0.17 |  |
|  | Provider | $41,101,881 | $19,495,413 | $3,505,541 | $1,273,358 | $2,583,229 | $8,978,966 | $9,245,330 | $86,183,718 | $3.33 | $0.55 |  |
|  | **Total** | $41,101,881 | $19,495,413 | $3,505,541 | $1,273,358 | $2,583,229 | $8,978,966 | $35,932,606 | $112,870,994 | $4.36 | $0.73 | $0.45 |
| Ghana | Household | $0 | $0 | $0 | $0 | $0 | $0 | $4,494,496 | $4,494,496 | $1.28 | $0.21 |  |
|  | Provider | $5,593,435 | $2,795,066 | $698,916 | $201,764 | $400,670 | $1,337,012 | $2,501,300 | $13,528,163 | $3.84 | $0.64 |  |
|  | **Total** | $5,593,435 | $2,795,066 | $698,916 | $201,764 | $400,670 | $1,337,012 | $6,995,795 | $18,022,659 | $5.12 | $0.85 | $0.57 |
| Niger | Household | $0 | $0 | $0 | $0 | $0 | $0 | $917,787 | $917,787 | $0.31 | $0.05 |  |
|  | Provider | $4,675,462 | $1,399,839 | $256,793 | $69,648 | $140,003 | $722,567 | $1,697,399 | $8,961,711 | $3.04 | $0.51 |  |
|  | **Total** | $4,675,462 | $1,399,839 | $256,793 | $69,648 | $140,003 | $722,567 | $2,615,186 | $9,879,498 | $3.35 | $0.56 | $0.28 |
| Cameroon | Household | $0 | $0 | $0 | $0 | $0 | $0 | $3,478,721 | $3,478,721 | $0.91 | $0.15 |  |
|  | Provider | $6,085,520 | $5,288,439 | $778,078 | $300,078 | $616,530 | $1,771,417 | $3,171,179 | $18,011,242 | $4.70 | $0.78 |  |
|  | **Total** | $6,085,520 | $5,288,439 | $778,078 | $300,078 | $616,530 | $1,771,417 | $6,649,900 | $21,489,963 | $5.61 | $0.93 | $0.65 |
| Burkina Faso | Household | $0 | $0 | $0 | $0 | $0 | $0 | $916,416 | $916,416 | $0.45 | $0.08 |  |
|  | Provider | $3,218,419 | $1,981,412 | $263,498 | $78,228 | $158,858 | $617,616 | $1,755,914 | $8,073,944 | $3.98 | $0.66 |  |
|  | **Total** | $3,218,419 | $1,981,412 | $263,498 | $78,228 | $158,858 | $617,616 | $2,672,330 | $8,990,360 | $4.43 | $0.74 | $0.46 |
| Mali | Household | $0 | $0 | $0 | $0 | $0 | $0 | $849,471 | $849,471 | $0.43 | $0.07 |  |
|  | Provider | $3,113,021 | $1,306,563 | $347,378 | $79,361 | $154,079 | $613,587 | $1,739,842 | $7,353,830 | $3.75 | $0.63 |  |
|  | **Total** | $3,113,021 | $1,306,563 | $347,378 | $79,361 | $154,079 | $613,587 | $2,589,313 | $8,203,301 | $4.18 | $0.70 | $0.42 |
| DRC | Household | $0 | $0 | $0 | $0 | $0 | $0 | $5,117,324 | $5,117,324 | $0.27 | $0.04 |  |
|  | Provider | $30,413,911 | $8,991,344 | $2,064,455 | $740,232 | $1,501,312 | $5,840,754 | $5,902,966 | $55,454,974 | $2.89 | $0.48 |  |
|  | **Total** | $30,413,911 | $8,991,344 | $2,064,455 | $740,232 | $1,501,312 | $5,840,754 | $11,020,291 | $60,572,298 | $3.16 | $0.53 | $0.25 |
| Mozambique | Household | $0 | $0 | $0 | $0 | $0 | $0 | $2,022,792 | $2,022,792 | $0.26 | $0.04 |  |
|  | Provider | $12,257,835 | $4,799,468 | $762,176 | $270,866 | $551,725 | $2,244,250 | $2,969,955 | $23,856,276 | $3.09 | $0.51 |  |
|  | **Total** | $12,257,835 | $4,799,468 | $762,176 | $270,866 | $551,725 | $2,244,250 | $4,992,747 | $25,879,068 | $3.35 | $0.56 | $0.28 |
| Tanzania | Household | $0 | $0 | $0 | $0 | $0 | $0 | $7,482,602 | $7,482,602 | $0.67 | $0.22 |  |
|  | Provider | $45,906,607 | $6,343,783 | $653,385 | $378,229 | $769,484 | $5,765,224 | $3,642,652 | $63,459,364 | $5.72 | $1.91 |  |
|  | **Total** | $45,906,607 | $6,343,783 | $653,385 | $378,229 | $769,484 | $5,765,224 | $11,125,254 | $70,941,966 | $6.40 | $2.13 | $0.67 |
| Uganda | Household | $0 | $0 | $0 | $0 | $0 | $0 | $4,153,823 | $4,153,823 | $0.52 | $0.09 |  |
|  | Provider | $66,057,657 | $4,087,075 | $751,512 | $264,305 | $537,792 | $7,342,082 | $2,926,797 | $81,967,220 | $10.27 | $1.71 |  |
|  | **Total** | $66,057,657 | $4,087,075 | $751,512 | $264,305 | $537,792 | $7,342,082 | $7,080,620 | $86,121,043 | $10.79 | $1.80 | $0.34 |

#### Table S6 Incremental cost-effectiveness ratios and learning adjusted years of school gained per $100 invested

*All costs presented in 2023 USD. ICERs are estimated using the provider perspective. Educational gains estimated using the foundational skills in literacy learning adjusted years of schooling (LAYS) approach. DALY: Disability-adjusted life year; DRC: Democratic Republic of the Congo; HBHI: High-burden high-impact; ICER: incremental cost-effectiveness ratio; LAYS: learning adjusted years of schooling*

|  | **Burkina Faso** | **Cameroon** | **DRC** | **Ghana** | **Mali** | **Mozambique** | **Niger** | **Nigeria** | **Tanzania** | **Uganda** | **All HBHI Countries** |
| --- | --- | --- | --- | --- | --- | --- | --- | --- | --- | --- | --- |
| **Intervention Drug & # of doses** | **SPAQ X6** | **SPAQ X6** | **SPAQ X6** | **SPAQ X6** | **SPAQ X6** | **SPAQ X6** | **SPAQ X6** | **SPAQ X6** | **DP X3** | **DP X6** |  |
| **Intervention Cost (millions)** | $8.1 | $18.0 | $55.5 | $13.5 | $7.4 | $23.9 | $9.0 | $86.1 | $63.5 | $82.0 | $366.9 |
| **Incremental Malaria Cases averted** | 265,002 | 386,411 | 2,169,909 | 297,961 | 256,515 | 825,922 | 337,742 | 2,995,407 | 366,632 | 1,217,059 | 9,118,561 |
| **Incremental DALYs averted** | 10,054 | 11,662 | 84,149 | 10,034 | 8,496 | 29,817 | 28,278 | 155,341 | 20,910 | 29,989 | 388,731 |
| **Incremental Morbidity Cost Savings** | $848,577 | $491,076 | $2,070,678 | $2,085,418 | $325,354 | $2,682,577 | $574,961 | $13,328,518 | $1,254,586 | $2,933,373 | $26,595,118 |
| **ICER (cost per case averted)** | $30 | $47 | $26 | $45 | $29 | $29 | $27 | $29 | $173 | $67 | $40 |
| **ICER (cost per DALY averted)** | $719 | $1,502 | $634 | $1,140 | $827 | $710 | $297 | $469 | $2,975 | $2,635 | $944 |
| **Incremental LAYS gains** | 434,523 | 821,614 | 4,106,224 | 755,177 | 420,293 | 1,654,947 | 631,240 | 5,549,221 | 2,377,282 | 1,710,404 | 18,460,926 |
| **Incremental Human Capital Gains due to Education (millions)** | $1,175 | $4,016 | $20,414 | $6,942 | $679 | $3,309 | $1,003 | $31,166 | $14,370 | $14,338 | $97,414,505,248 |
| **LAYS gained per $100 spent** | 5.38 | 4.56 | 7.40 | 5.58 | 5.72 | 6.94 | 7.04 | 6.44 | 3.75 | 2.09 | 5.03 |

### Sensitivity Analysis

#### Figure S1 Tornado Diagram for benefit-cost ratio

*The tornado diagram illustrates the correlation between the simulated generalized benefit-cost ration and the sampled input values, using the absolute values of these correlations to highlight influential parameters. IPTsc = intermittent preventative treatment in school-age children; OOP = out of pocket payments ; SAC = school-age children*

|  | Absolute value of correlation |
| --- | --- |
| OOP for transportation and food for uncomplicated malaria | 0.00126314 |
| Number of days lost school due to one episode of uncomplicated malaria | 0.00133099 |
| Duration of hosital stay for severe malaria | -0.00144318 |
| Number of days lost school due to one episode of severe malaria | 0.00189961 |
| Percentage of cases that will become severe malaria in SAC | 0.00545867 |
| Number of days of caregiver lost productivity for uncomplicated malaria | 0.00569674 |
| OOP of transportation and food for severe malaria | 0.00724329 |
| Percentage of care sought at publicly funded health centers | -0.00904838 |
| Cost of hospitalization per day in public facility | 0.00971479 |
| Number of days of caregiver lost productivity for untreated malaria | 0.00972506 |
| Number of days of caregiver lost productivity for severe malaria | 0.0105364 |
| Cost per outpatient consultation fee | 0.012654 |
| Estimated returns to schooling | 0.0148067 |
| Number of teacher-hours used to develiver IPTsc per round per school | -0.0171409 |
| Cost of freight and insurance | -0.0172123 |
| Cost of wastage | -0.0218797 |
| Probablity of seeking care | -0.0259607 |
| Percentage of parents that attend community sensitization meeting | -0.0398829 |
| Intervention coverage | -0.0401178 |
| Monthly teacher salary | -0.0764202 |
| Value of day of lost productivity to households | -0.112673 |
| Rate ratio effect size of IPTsc | -0.394447 |
| Incidence rate | 0.414525 |
| Case fatality rate | 0.772597 |

#### Table S7 Drug choice sensitivity analysis

To illustrate the impact of drug choice on both costs and benefits, we modeled two scenarios for Uganda: one using dihydroartemisinin-piperaquine (DP) and one using sulfadoxine-pyrimethamine plus amodiaquine (SPAQ), each administered six times per year. Uganda was selected because a trial of malaria chemoprevention in school-aged children (SAC) is planned there, and it is plausible that either drug could be used both in the trial and in future programmatic implementation.

In this analysis, drug price (DP: $1.46 per dose; SPAQ: $0.28 per dose) and effect size (DP incidence rate ratio [IRR]: 0.50; SPAQ IRR: 0.665) were the only parameters varied. The resulting annual cost per pupil was $10.79 for DP and $3.76 for SPAQ. Despite DP’s greater protective effect, SPAQ yielded a higher benefit-cost ratio (BCR), with a BCR of 3.71 for DP and 7.74 for SPAQ. These findings highlight the importance of drug choice in determining both the cost and value for money of malaria chemoprevention programs.

| **Societal perspective** |  |  |
| --- | --- | --- |
| Intervention drug | **DP** | **SPAQ** |
| Intervention cost | $86,121,043 | $27,647,305 |
| Incremental Malaria Cases averted | 1,217,059 | 815,430 |
| Incremental Deaths averted | 1,544 | 1,034 |
| Incremental morbidity cost savings | $27,810,495 | $18,633,032 |
| Incremental mortality cost savings | $269,078,856 | $180,282,834 |
| Net Benefits (health only) | $210,768,308 | $171,268,561 |
| Benefit-cost ratio (health only) | 3.45 | 7.19 |
| Incremental school years gains | 16,963 | 11,365 |
| Incremental Human Capital Gains due to Education | $22,412,983 | $15,016,699 |
| Net Benefits (health and education) | $233,181,291 | $186,285,260 |
| Benefit-cost ratio (health and education) | 3.71 | 7.74 |
